# Severe COVID-19 infection is associated with aberrant cytokine production by infected lung epithelial cells rather than by systemic immune dysfunction

**DOI:** 10.1101/2021.12.09.21266492

**Authors:** Sherin J Rouhani, Jonathan A Trujillo, Athalia R Pyzer, Jovian Yu, Jessica Fessler, Alexandra Cabanov, Emily F Higgs, Kyle R. Cron, Yuanyuan Zha, Yihao Lu, Jeffrey C. Bloodworth, Mustafa Fatih Abasiyanik, Susan Okrah, Blake A Flood, Ken Hatogai, Michael YK Leung, Apameh Pezeshk, Lara Kozloff, Robin Reschke, Garth W. Strohbehn, Carolina Soto Chervin, Madan Kumar, Stephen Schrantz, Maria Lucia Madariaga, Kathleen G Beavis, Kiang-Teck J. Yeo, Randy F. Sweis, Jeremy Segal, Savaş Tay, Evgeny Izumchenko, Jeffrey Mueller, Lin S Chen, Thomas F Gajewski

## Abstract

The mechanisms explaining progression to severe COVID-19 remain poorly understood. It has been proposed that immune system dysregulation/over-stimulation may be implicated, but it is not clear how such processes would lead to respiratory failure. We performed comprehensive multiparameter immune monitoring in a tightly controlled cohort of 128 COVID-19 patients, and used the ratio of oxygen saturation to fraction of inspired oxygen (SpO2 / FiO2) as a physiologic measure of disease severity. Machine learning algorithms integrating 139 parameters identified IL-6 and CCL2 as two factors predictive of severe disease, consistent with the therapeutic benefit observed with anti-IL6-R antibody treatment. However, transcripts encoding these cytokines were not detected among circulating immune cells. Rather, in situ analysis of lung specimens using RNAscope and immunofluorescent staining revealed that elevated IL-6 and CCL2 were dominantly produced by infected lung type II pneumocytes. Severe disease was not associated with higher viral load, deficient antibody responses, or dysfunctional T cell responses. These results refine our understanding of severe COVID-19 pathophysiology, indicating that aberrant cytokine production by infected lung epithelial cells is a major driver of immunopathology. We propose that these factors cause local immune regulation towards the benefit of the virus.

## Introduction

The clinical manifestations of COVID-19 range in severity from asymptomatic infection to critical illness and death, yet the mechanisms by which SARS-CoV-2 cause morbidity and mortality have yet to be fully elucidated. It has been proposed that an excessive immune response may cause immunopathology in affected target organs, particularly the lower respiratory tract. Several large studies of hospitalized patients demonstrated that disease severity and mortality are correlated with elevated levels of inflammatory cytokines, suggesting a potentially dysregulated immune response to infection^1–4^. Consistent with this notion, the steroid dexamethasone improved outcomes in severe and critically ill patients^5,6^. IL-6 specifically has been proposed as a functionally important cytokine,^7^ and the anti-IL-6R antibody (Ab) tocilizumab provided a survival benefit in critically ill COVID-19 patients^8,9^.

SARS-CoV-2-infected patients can develop both T cell and B cell responses^2,3,10–12^. Some groups of patients appear to develop phenotypically distinct immune responses, which have been hypothesized to be maladaptive^2–4,10,13^. This includes skewing towards a Th2 or Th17 phenotype^2^ or uncoordinated B and/or T cell responses^10^. However, these studies used heterogeneous cohorts of patients at different phases of infection, and concurrent disease states or immunosuppression may complicate the interpretation of immunologic studies. African American and Latino patients are disproportionately affected by the SARS-CoV-2 pandemic, but they generally are under-represented in translational research studies. Similarly, analysis of non-hospitalized patients with mild COVID-19 has been limited. Based on these considerations, we examined the longitudinal immune response from non-immunosuppressed, predominantly African American and Latino patients. We defined disease severity based on the ratio of oxygen saturation to fraction of inspired oxygen (SpO2 / FiO2). Immune parameters associated with disease severity were identified based on an unbiased machine learning algorithm. When a disconnect was identified between elevated serum cytokine levels yet lack of evidence for their production by immune cells, lung tissue was studied for in situ expression of key immune genes.

## Results

### Patients and definition of disease severity

We analyzed 101 hospitalized COVID-19 patients, 27 non-hospitalized COVID-19 outpatients, and 22 healthy donors (HD) as part of a COVID-19 biobanking protocol (Fig 1a). Sixty-seven additional COVID-19 patients were excluded from analysis because of potential immunological confounders, as listed in Supplementary Table 1. Samples from patients who received the anti-IL-6R antibody tocilizumab were excluded from cytokine analyses, as tocilizumab can modulate levels of IL-6 and other cytokines^14^. Demographic characteristics of the study cohort are shown in Supplementary Table 2. Our patient population was 68% African-American with a median age of 55 years. To avoid over-sampling bias from severe patients who had more timepoints available for analysis, we used the maximum level of soluble factors quantified per patient from an early (Day 1-9) and late (Day 10-30) timepoint post-symptom onset, except when assessing cytokine kinetics.

**Figure 1:**
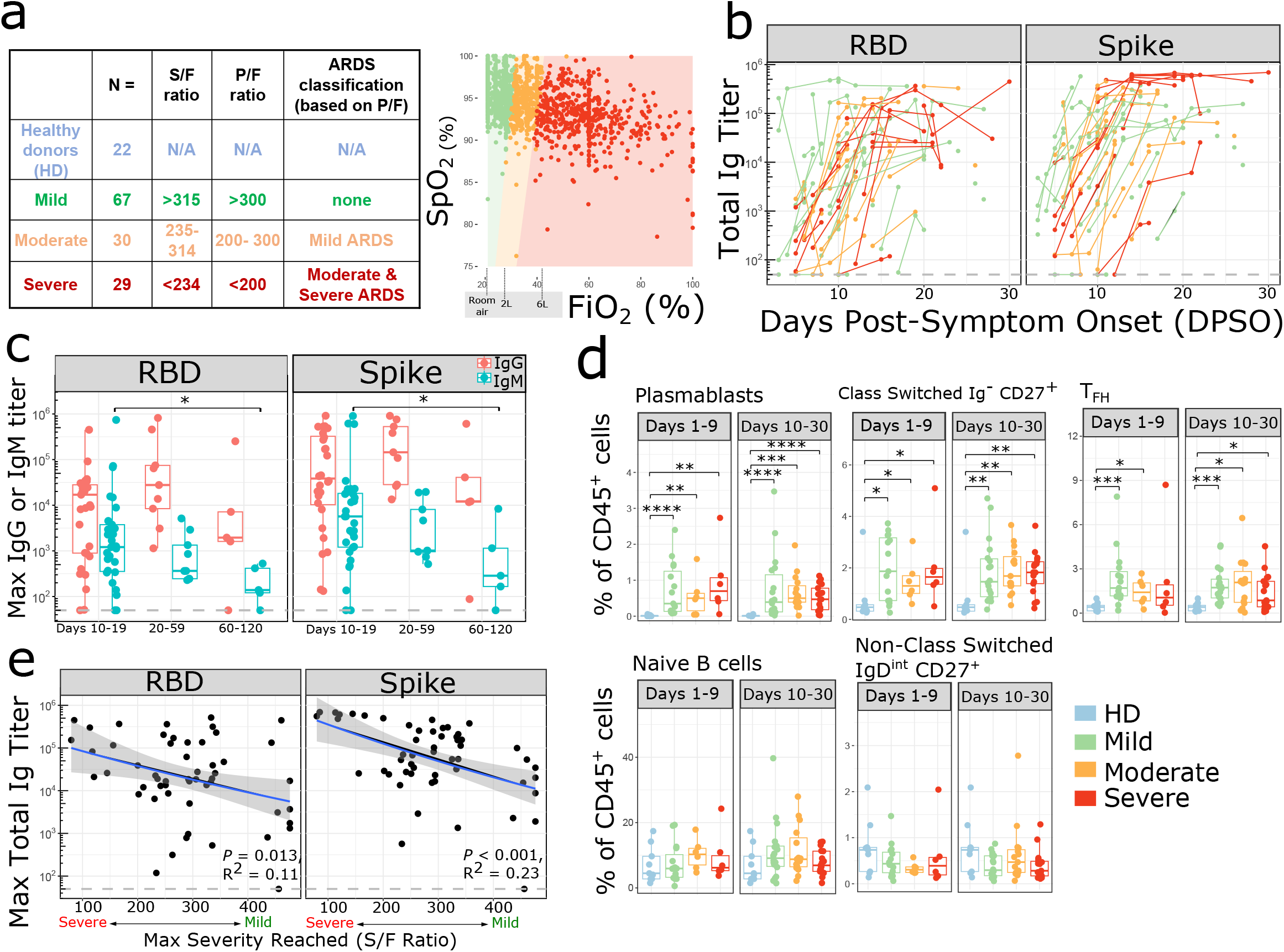
Activated B cells and antibody responses are induced within 10 days of SARS-CoV-2 symptom onset (a) Number (N) of patients in each disease severity classification. The table describes the relationship between SaO2/FiO2 (S/F), PaO2/FiO2 (P/F) ratio and ARDS classes for mild, moderate and severe categories based on Rice et al15. The relationship between SpO2 and FiO2 is depicted on the right. Each dot represents an individual S/F ratio calculated based on the simultaneous oxygen saturation and FiO2 recorded for hospitalized patients; all available timepoints per day per patient are shown. Oxygen delivery categories are labelled on the X axis. Colored shaded areas indicate S/F ratios which correspond to mild (green), moderate (orange) and severe (red) disease severity categories. (b) Serum SARS-CoV-2 total Ig antibody levels (RBD and Spike) in (n=68) PCR+ patients over time expressed as days post symptom onset (DPSO). Samples from individual patients are connected by lines, and colored by disease severity (mild in green, moderate in orange, severe in red). The dashed line indicates the max titer of a pool of negative controls, used as the threshold for positivity. Samples post receipt of convalescent plasma transfusion were excluded from this and all antibody analyses. (c) Maximum serum SARS-CoV-2 IgG (red) and IgM (turquoise) antibody levels (RBD and Spike) in PCR+ patients during acute phase of infection (day 10-19) (n=31), recovery (day 20-59) (n=9), and late recovery (day 60-120) (n=5), where day is DPSO. Data from the pre-humoral phase (<day 10) is excluded. (d) Mean populations of immune cell subsets involved in the humoral response, plotted as % of CD45+ cells, during the early phase (days 1-9 from symptom onset) and late phase (days 10-30 from symptom onset) of disease in COVID-19 infection compared to non-infected healthy donor controls (HD). Each dot represents the average of measurements during each phase of disease from an individual subject (early phase: mild, n=15; moderate, n=6; severe disease, n= 6 and late phase: mild, n=18; moderate, n=15; severe, n=17; non-infected healthy controls, n=9). The boxplots show the medians (middle line) and the first and third quartiles (upper and lower bounds of the boxes). (c,d) Significance was determined by two-sided Mann Whitney Wilcoxon test and p-values are indicated by asterisks (*, p ≤ 0.05; **, p ≤ 0.01; ***, p ≤ 0.001, ****, p ≤ 0.0001). (e) Linear regression shown for disease severity expressed as S/F ratio and maximum anti-SARS-CoV-2 (RBD and Spike) total Ig antibody titers from days 10-30 in (n=53) PCR+ patients. Shaded areas represent 95% confidence interval. Ig titers from the pre-humoral phase (<day 10) were excluded.

To obtain an objective measure of disease severity to correlate with immunologic parameters, the pulse oximeter oxygen saturation (SpO_2_) / fraction of inspired oxygen (FiO_2_) ratio (S/F ratio) was calculated for each patient over time (Fig 1a). The S/F ratio is analogous to the PaO_2_/FiO_2_ (P/F) ratio used in ARDS studies^15,16^ and has been validated as an independent correlate of severity in SARS-CoV-2 infection^16^. Patients were divided into three groups - mild, moderate, and severe - on the basis of their worst daily mean S/F ratio during their initial hospitalization. Patients with normal oxygen saturation on room air and outpatients were defined as mild (S/F > 315), while the majority of inpatients on non-invasive or invasive ventilatory support were classified as severe (Supplementary Fig 1a). The S/F ratio is a dynamic and objective measurement of a patient’s respiratory status over the course of illness and hospitalization (Supplementary Fig 1c) and provides a continuous scale of disease severity.

### Robust adaptive immune responses in infected patients

Consistent with other studies^17^, severe patients had higher maximum C-reactive protein (CRP), ferritin, and D-dimer levels compared to mild patients (Supplementary Fig 1d). The absolute lymphocyte count decreased with worsened disease severity, and many patients were lymphopenic.

One hypothesis potentially explaining disease severity was a diminished or delayed adaptive immune response, leading to failed viral clearance. We therefore measured total immunoglobulin (Ig), IgG, and IgM antibody titers against the SARS-CoV-2 Spike glycoprotein and its Receptor Binding Domain (RBD). RBD binds to angiotensin-converting enzyme 2 (ACE2) receptor on human cells and is the primary target for neutralizing antibodies^18–20^. By day 10 post-symptom onset, 44 of 45 evaluable patients had detectable anti-Spike and anti-RBD total Ig (Figure 1b). IgG titers to Spike and RBD persisted through the acute phase of infection (day 10-19), recovery (day 20-59), and into late recovery (day 60 – 120), whereas IgM titers started to decline in the recovery phase, as expected (Figure 1c). There was a corresponding increase in the frequency of antibody-producing plasmablasts, class switched IgD^neg^ B cells, and T follicular helper cells (Tfh) by day 9 (Fig 1d). When examining Ab responses by disease severity, patients with severe disease developed comparable maximum anti-RBD and anti-Spike antibody titers compared to patients with mild or moderate disease (Figure 1b, 1e), indicating that a failed Ab response was not causal for progression to severe disease. Spike and RBD titers also did not correlate with age (Supplementary Fig 2a) or gender (Supplementary Fig 2b). IL-6 is known to be involved in plasma cell differentiation and antibody production^21,22^, so we investigated whether treatment with the IL-6R antagonist tocilizumab affected SARS-CoV-2 antibody generation; yet no differences were observed (Supplementary Fig 2c). Anti-viral medications such as remdesivir could have decreased antigen load and led to a lesser Ab response; however, no diminution of Ab response was observed (Supplementary Fig 2c). Consistent with these results, nasopharyngeal viral load as measured by digital droplet PCR (ddPCR) did not differ between patients with mild, moderate, or severe disease (Supplementary Fig 2d).

Analysis of circulating T cells by flow cytometry (Supplementary Fig 3) revealed a decrease in the percentage of CD8^+^ T cells relative to total CD45^+^ cells in severe patients (Fig 2a). The percentages of CD8^+^ central memory (CM) and effector memory (EM) cells decreased at both early and late time points, while the frequency of terminally differentiated memory (TEMRA) cells was relatively stable (Fig 2a). The percentage of CD4^+^ CM cells increased in patients while the percentage of CD4^+^ EM cells remained unchanged. The percentage of T helper type 1 (Th1) cells was increased in severe COVID-19 patients while the percentage of T helper type 2 (Th2) cells did not change. CD4^+^ T cells upregulated CD57, a marker of cytotoxic terminally differentiated cells,^23,24^ and CD8^+^ T cells upregulated CD95 (Supplementary Fig 4a,4b).

**Figure 2:**
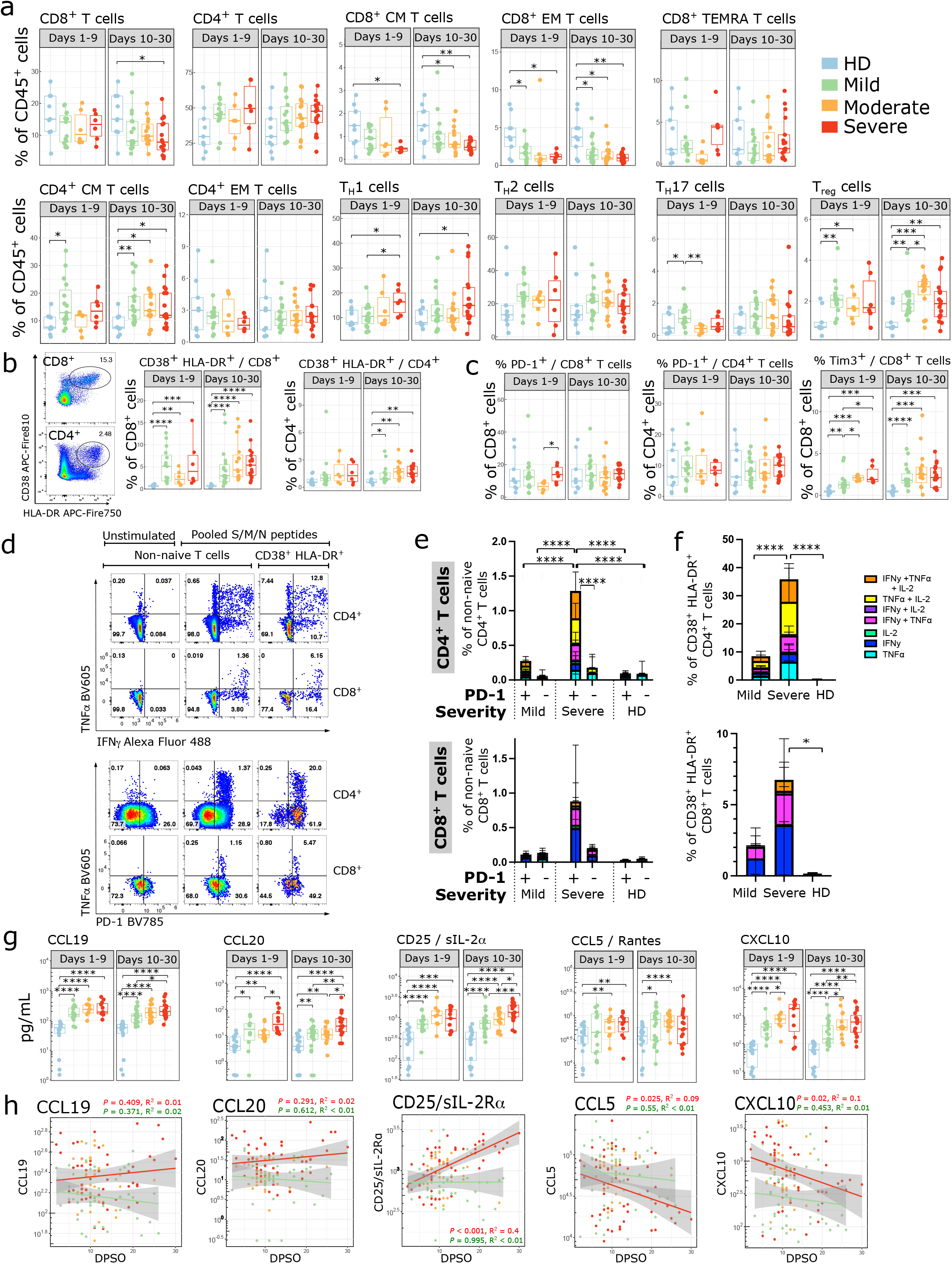
SARS-CoV-2 infection elicits a robust expansion of activated polyfunctional T cells (a-c) Proportion of immune cell subsets related to adaptive immune responses are shown as a percentage of either live CD45+ cells, live CD4+ cells, or live CD8+ cells, as indicated. Where multiple timepoints within the early (D1-9) or late (D10-30) phase per patient were available, the mean was taken and each patient is represented by one dot per time phase. n = 9 for HD, n = 15, 6, 6 for mild, moderate, and severe respectively in the early phase, and n = 18, 15, 17 for mild, moderate, and severe in the late phase. (b, left) Representative flow cytometry plots showing CD38+ HLA-DR+ subsets. (d-f) PBMCs from D11-25 DPSO were stimulated with a combined pool of peptides from the S, M and N proteins for 9 hours and stained for intracellular cytokine production. Background activity in unstimulated wells was subtracted from stimulated wells; negative values after subtraction were set to 0. Representative flow cytometry plots are shown for TNF**α**/IFN-γ (d, upper panels) and TNF**α**/PD-1 (d, lower panels) staining. The percentage of cells producing various combinations of IFN-γ, TNF**α**, and IL-2 were reported for CD4+ and CD8+ non-naive (e) and CD4+ and CD8+ CD38+HLADR+ cells (f). (e,f) Comparisons between groups were done by summing the total cytokine production in each column and performing a 2-way ANOVA with Sidak’s multiple comparisons test. Error bars represent mean +/- SD for each cytokine subset. HD n = 9; mild n = 8; severe n = 7. Wells with <50 CD38+HLA-DR+ cells were excluded from that subset analysis, leaving n = 9 / 7 / 6 HD/mild/severe for CD4+CD38+HLA-DR+ and n = 9 / 8 / 6 HD/mild/severe for CD8+CD38+HLA-DR+. (g) Peak cytokine and chemokine levels related to T cell activation and survival are shown during the early phase (days 1-9 from symptom onset) and late phase (days 10-30 from symptom onset) of disease in SARS-CoV-2 infection compared to non-infected healthy controls. Each dot represents maximum value per individual subject during each phase of disease (early phase: mild, n=15; moderate, n=10; severe, n= 11 and late phase: mild, n=23; moderate, n=16; severe, n= 19 and non-infected healthy controls, n=18). Samples from patients post receipt of tocilizumab (which directly modulates cytokine levels) were excluded from this and subsequent cytokine analyses. (h) Kinetics of cytokine expression over time (days post symptom onset) from mild (green, n = 33), moderate (orange, n = 19), and severe (red, n = 23) patients. Multiple timepoints per patient plotted when available. Linear regression for cytokine values over time in severe (red) and mild (green) patients shown. Shaded areas represent 95% confidence interval. (a-c, g) Significance was determined by two-sided Mann Whitney Wilcoxon test and p-values are indicated by asterisks (*, p ≤0.05; **, p ≤ 0.01; ***, p ≤ 0.001, ****, p ≤ 0.0001).

CD38 and HLA-DR are markers of activated T cells during viral infections^25^, and this population was increased among both CD4^+^ and CD8^+^ T cells in COVID-19 patients of all disease severities (Fig 2b). It was particularly striking in CD8^+^ T cells, where 42% (21 / 50) of patients had > 5% of all CD8^+^ T cells expressing these activation markers between days 10-30. Thus, despite a relative overall lymphopenia, there was an abundance of activated T cells in severe patients. Self-clustering analysis using UMAP and FlowSOM algorithms^26^ showed COVID-19 patients had increased percentages of activated CD8^+^ (cluster 4) and CD4^+^ (cluster 11) cells expressing high levels of CD38, HLA-DR, and CD95 (Supplementary Fig 4c-e). CD8^+^ CM cells and multiple subsets of CD4^+^ T cells upregulated CD28 (Supplementary Fig 4a,4b). There were no significant differences in the percentages of CD4^+^ or CD8^+^ T cells expressing PD-1, although modest upregulation of PD-1 was seen on CD4^+^ EM and CD8^+^ EM and TEMRA cells (Fig 2c, Supplementary Fig 4a,b). There was an increased percentage of CD8^+^ T cells expressing TIM-3 (Fig 2c). The proportion of regulatory T cells (Tregs) also increased in COVID-19 patients (Fig 2a), suggesting a counter-regulatory mechanism in response to increasing levels of T cell activation.

Several studies have shown that inhibitory receptors including PD-1 are upregulated on SARS-CoV-2 specific T cells, and have suggested that PD-1^high^ cells in COVID-19 infection are exhausted^17,27–29^ or have decreased polyfunctionality^28,30^. However, PD-1 can also be upregulated in acutely activated T cells^3,31^. To determine whether there were differences in IFN-γ production by SARS-CoV-2-specific T cells, we used an ELISPOT to measure IFN-γ production after stimulation with overlapping HLA class I & II 15-mer peptides from the S, M, and N proteins of SARS-CoV-2. IFN-γ production was seen as early as day 8 after symptom onset, and the degree of IFN-γ production was similar between patients with different disease severities (Supplementary Fig 4f).

To determine if PD-1 on these cells represents a marker of activation or exhaustion, we used intracellular cytokine staining to measure polyfunctionality after S/M/N peptide stimulation. Compared to mild patients, severe patients had higher percentages of polyfunctional CD4^+^ T cells producing IFN-γ, TNF-α, and/or IL-2 in response to S/M/N peptide stimulation (Fig 2d-f). Furthermore, cytokine production was concentrated in the PD-1^+^CD4^+^ T cells, indicating that PD-1 represents an activation marker rather than a marker of dysfunction in this context. There was a similar trend with PD-1^+^ CD8^+^ T cells in severe patients, but this was not significant due to increased patient-to-patient variability in the CD8^+^ T cell response (Supplementary Fig 4g). Cytokine-producing T cells were enriched amongst the CD38^+^HLA-DR^+^ population (Fig 2d,2f), consistent with this population containing virus-activated T cells. We conclude that the adaptive immune response is robust in severe COVID-19 patients and that lack of virus-specific immunity is not contributory to the progression to disease severity.

Patients with SARS-CoV-2 also had a serum cytokine and chemokine profile consistent with increased T cell activation. Levels of CCL19 and CCL20, which recruit T cells to lymph nodes for activation, increased with disease severity (Fig 2g). Severe patients had higher levels of sCD25/IL-2Ra, which is cleaved and released upon T cell activation. CCL5 and CXCL10 recruit T cells to sites of inflammation, and were elevated in the serum of COVID-19 patients. CXCL10 also increased with disease severity. Levels of CCL19, CCL20, and CD25/IL-2Ra remained elevated over time in severe patients, while CCL5 and CXCL10 levels declined over time in both mild and severe patients (Fig 2h). Patients with severe disease had increased levels of T cell survival cytokines IL-15 and IL-7 (Supplementary Fig 4h). Levels of the immunoregulatory molecules IL-10 and IL-1RA were increased in patients with SARS-CoV-2, suggestive of an expected negative feedback loop in response to increasing T cell activation^32,33^.

### Innate immune cells and circulating cytokines

Analysis of the innate immune response demonstrated a decreased proportion of circulating NK cells and particularly the CD16^-^ NK cell subset at early and late timepoints (Fig 3a). Frequencies of dendritic cell (DC) subsets remained mostly unchanged other than a decrease in CD123^+^ plasmacytoid dendritic cells at late timepoints (Supplementary Fig 5a). However, the level of CD86 increased in plasmacytoid DCs, indicating a more activated status. CD1c^+^ DCs also had higher levels of Tim-3 at late time points after SARS-CoV-2 infection. The proportions of neutrophils, non-classical monocytes, and intermediate monocytes were increased in patients with COVID-19 compared to healthy controls (Fig 3a). While the percentage of classical monocytes was unchanged, the mean fluorescence intensity (MFI) of CD86 and HLA-DR was decreased in infected patients (Fig 3a), suggesting the emergence of less-mature monocytes from the bone marrow. This is further supported by increased levels of the myeloid growth factor GM-CSF in the serum of patients with SARS-CoV-2 infection (Fig 3b), and a negative correlation between GM-CSF and HLA-DR levels on intermediate monocytes (R^2^ = 0.15, p = 0.004) (Supplementary Fig 5c). These parameters are consistent with tissue repair-type macrophages being favored during SARS-CoV-2 infection.

**Figure 3:**
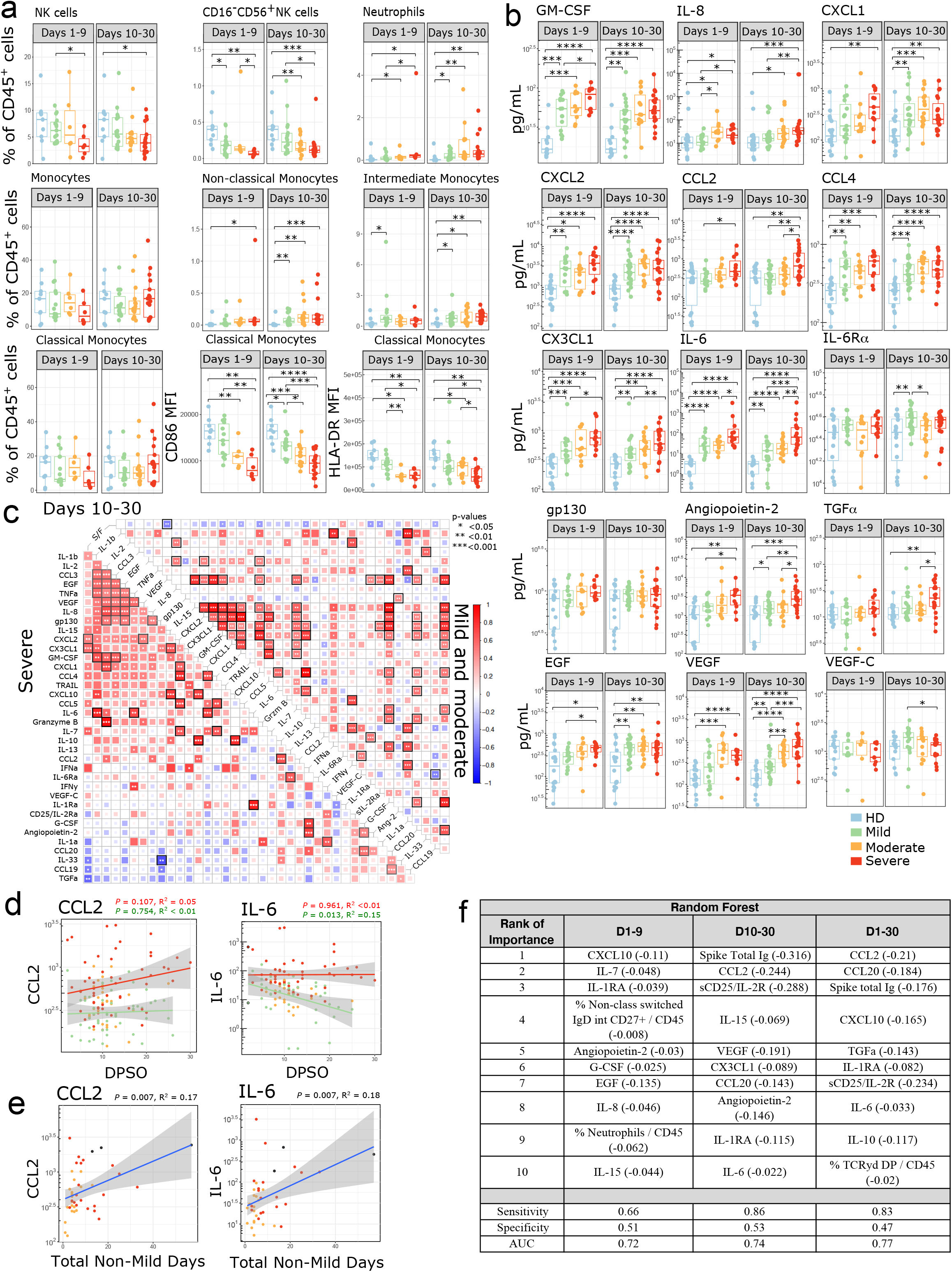
Innate immune changes in SARS-CoV-2 infection (a) Immune cell subsets related to the innate immune response are shown. Boxplots show percentages of each cell population out of live CD45+ cells or MFI of indicated markers. Where multiple timepoints within the early (D1-9) or late (D10-30) phase per patient were available, the mean was taken and each patient is represented by one dot per time phase. n = 9 for HD, n = 15, 6, 6 for mild, moderate, and severe, respectively in the early phase, and n = 18, 15, 17 for mild, moderate and severe in the late phase. (b) Peak cytokine and chemokine levels are shown during the early phase (days 1-9 from symptom onset) and late phase (days 10-30 from symptom onset) of disease in SARS-CoV-2 infection compared to non-infected healthy controls. Each dot represents maximum value per individual subject during each phase of disease (early phase: mild, n=15; moderate, n=10; severe, n= 11 and late phase: mild, n=23; moderate, n=16; severe, n= 19 and non-infected healthy controls, n=18). (c) Correlations between cytokines from days 10-30 for COVID-19 patients were calculated and clustered hierarchically. S/F ratio is fixed as the first column for comparison. Samples were stratified by disease severity. Spearman correlation coefficients were quantified by the scale of color and size of colored squares; significance of the correlation is labeled with * (P < 0.05), ** (P < 0.01), and *** (P < 0.001). Black border represents a false-discovery rate (FDR) < 0.05 (d) Kinetics of cytokine expression over time (days post symptom onset) from mild (green, n = 33), moderate (orange, n = 19), and severe (red, n = 23) patients. Multiple timepoints per patient plotted when available. Linear regression for cytokine values over time in severe (red) and mild (green) patients shown. (e) Peak individual levels of CCL2 and IL-6 are shown as linear correlation with the sum of days each patient spent hospitalized with a moderate or severe S/F ratio, termed “total non-mild days”. Each dot represents maximum cytokine value per individual subject; maximal disease severity indicated by color (moderate [orange], n=20; severe [red], n= 19, deceased [black], n= 3). (d-e) Shaded areas represent 95% confidence interval. (f) The top 10 ranked immune parameters associated with severity per the Random Forest model are tabulated for early phase (days 1-9 from symptom onset) and late phase (days 10-30 from symptom onset) and all time-points (day 1-30 from symptom onset). The linear regression R2 value for each variable is shown in parenthesis, indicating the amount of variation in disease severity that can be explained by this variable alone. +/- denotes direction of association, + indicating the higher the variable the higher (i.e., less severe) the S/F ratio, and - indicating the higher the variable the lower the S/F ratio (i.e. the more severe the disease). Sensitivity, Specificity and AUC are shown. (a-b) Significance was determined by two-sided Mann Whitney Wilcoxon test and p-values are indicated by asterisks (*, p ≤ 0.05; **, p ≤ 0.01; ***, p ≤ 0.001, ****, p ≤ 0.0001).

Patients with SARS-CoV-2 had increased levels of cytokines responsible for recruiting neutrophils, monocytes and macrophages to sites of inflammation, including the neutrophil chemoattractants IL-8, CXCL1, CXCL2, and the monocyte chemoattractants CCL2, CCL4, and CX3CL1 (Fig 3b). IL-8, CCL2, and CX3CL1 also increased with disease severity. Distinct groups of cytokines clustered together in correlation plots at late time points (Fig 3c), particularly in severe patients. CCL2 levels remained high over time in severe patients, and higher levels of CCL2 also correlated with a longer duration of moderate or severe illness (R^2^ = 0.17, p = 0.00737) (Fig 3d-e).

IL-6 has been identified as a pathologic mediator of cytokine release syndrome after CAR-T cell treatment, and it has been hypothesized that a similar phenomenon may be driving severe pathology in some COVID-19 patients^7^. IL-6 signals through the IL-6R and gp130 complex. Gp130 is ubiquitously expressed, while IL-6R expression is normally limited to immune cells and hepatocytes. IL-6 can also form a complex with soluble IL-6R (sIL-6R**α**) and signal in trans through gp130 in cells that do not express the IL-6R. We found that IL-6 levels increased with disease severity, while sIL-6R**α** and gp130 levels were similar between severity groups (Fig 3b). While there was a correlation between CRP and IL-6 levels, there were many patients who had a high CRP but only a modest increase in IL-6 (Supplementary Fig 5d). Levels of IL-6 remained high at late timepoints in severe patients when compared to mild (Fig 3d), and levels of soluble gp130 were lower in severe patients at late timepoints (Supplementary Fig 5e). sIL-6R**α** levels remained high over time in both mild and severe patients. Interestingly, the duration of moderate or severe disease positively correlated with IL-6 levels and negatively correlated with soluble levels of gp130, which is an endogenous inhibitor of IL-6 trans-signaling ^34–37^ (Fig 3e, Supplementary Fig 5f).

In order to better understand the pathophysiology that differentiates severe patients from mild or moderate patients, we used the Random Forest machine learning algorithm with 3-fold cross-validation to model the impact of 139 defined immune parameters in an unbiased fashion. From the resulting model, the highest importance features were extracted (Figure 3f), and linear regression modeling was used to determine the relative impact of each feature. At early time points (D1-9), severe patients showed evidence for an active innate immune response (elevated G-CSF, IL-8, and the percentage of neutrophils) as well as an activated T cell response (elevated CXCL10, IL-7, and IL-15). Integrating the data from all phases (D1-30) of the immune response, a signature suggestive of T cell recruitment and activation with elevated CCL20, CXCL10, and sCD25/IL-2R was evident in severe patients, consistent with the notion that persistence of virus drives a continued T cell response. Additionally, severe patients had elevated levels of the macrophage related factors CCL2 and IL-6, with elevated CCL2 being the overall top-ranked immunological predictor of severe disease.

### IL-6 and CCL2 are produced by infected lung epithelial cells

Elevated levels of serum IL-6 and CCL2 were each associated with and predictive of severe COVID-19 disease. CCL2 is known to recruit macrophages, particularly M2 macrophages, into tissues. A detrimental role for IL-6 has been supported by studies showing improved clinical outcome upon treatment with the anti-IL-6R antibody tocilizumab^9^. Based on prior work studying cytokine-release syndrome in CAR-T cell therapy^38,39^ and IL-6 production in infectious models^40–42^, it has been assumed that IL-6 in COVID-19 patients is being produced by macrophages^43^. However, in our cohort examining representative patients having “ high” versus “ low” serum IL-6 levels at the protein level (Supplementary Fig 6a), no difference in mRNA for either IL-6 or CCL2 was observed among peripheral level blood mononuclear cells (Supplementary Fig 6c-d). This result is consistent with the flow cytometric analysis of circulating monocytes, which indicated an immature and possibly tissue repair phenotype rather than an inflammatory one (Fig 3a). Together, these results suggested that the source of these cytokines might not be immune cells, but rather raised the possibility that virus-infected cells in the lung might be the major source. We therefore examined expression of IL-6 and CCL2 mRNA in lung tissue from a cohort of 10 fatal COVID-19 cases listed in Supplementary Table 3. We performed a multispectral immunofluorescence assay combining RNA *in situ* hybridization (RNA-ISH) for SARS-CoV-2 RNA and IL-6 or CCL2 mRNA, along with protein immunofluorescence (IF) staining to identify the cells of origin. Thyroid transcription factor 1 (TTF1) was used to identify type 2 pneumocytes, and CD45 was utilized to identify leukocytes (Fig 4a, Supplementary Fig 7a). SARS-CoV-2 RNA was detected in all of the autopsy lung specimens. Unexpectedly, the vast majority of IL-6 transcripts were detected in cells that did not co-stain for the macrophage markers CD68 or the M2 macrophage marker CD163 (Supplementary Fig 6e-f). Interestingly, large numbers of TTF1^+^ type 2 pneumocytes expressed IL-6 mRNA, with a high percentage of these cells also positive for SARS-CoV-2 RNA (Fig 4a-c). Quantitative analysis showed TTF1^+^ type 2 pneumocytes were the predominant IL-6-expressing cell type, greatly outnumbering CD45^+^ immune cells (Fig 4b,c). Among the IL-6 positive populations, type 2 pneumocytes relative to CD45^+^ cells showed greater IL-6 expression on a per cell basis, as indicated by a greater number of TTF1^+^ cells with higher mean staining intensity for IL-6 (Fig 4d). Similarly, CCL2 expression was particularly abundant on TTF2^+^ type 2 pneumocytes (Supplementary Fig 7a-d). Together these data show that virus-infected lung epithelial cells are the major source of IL-6 and CCL2 in SARS-CoV-2 infected lungs.

**Figure 4:**
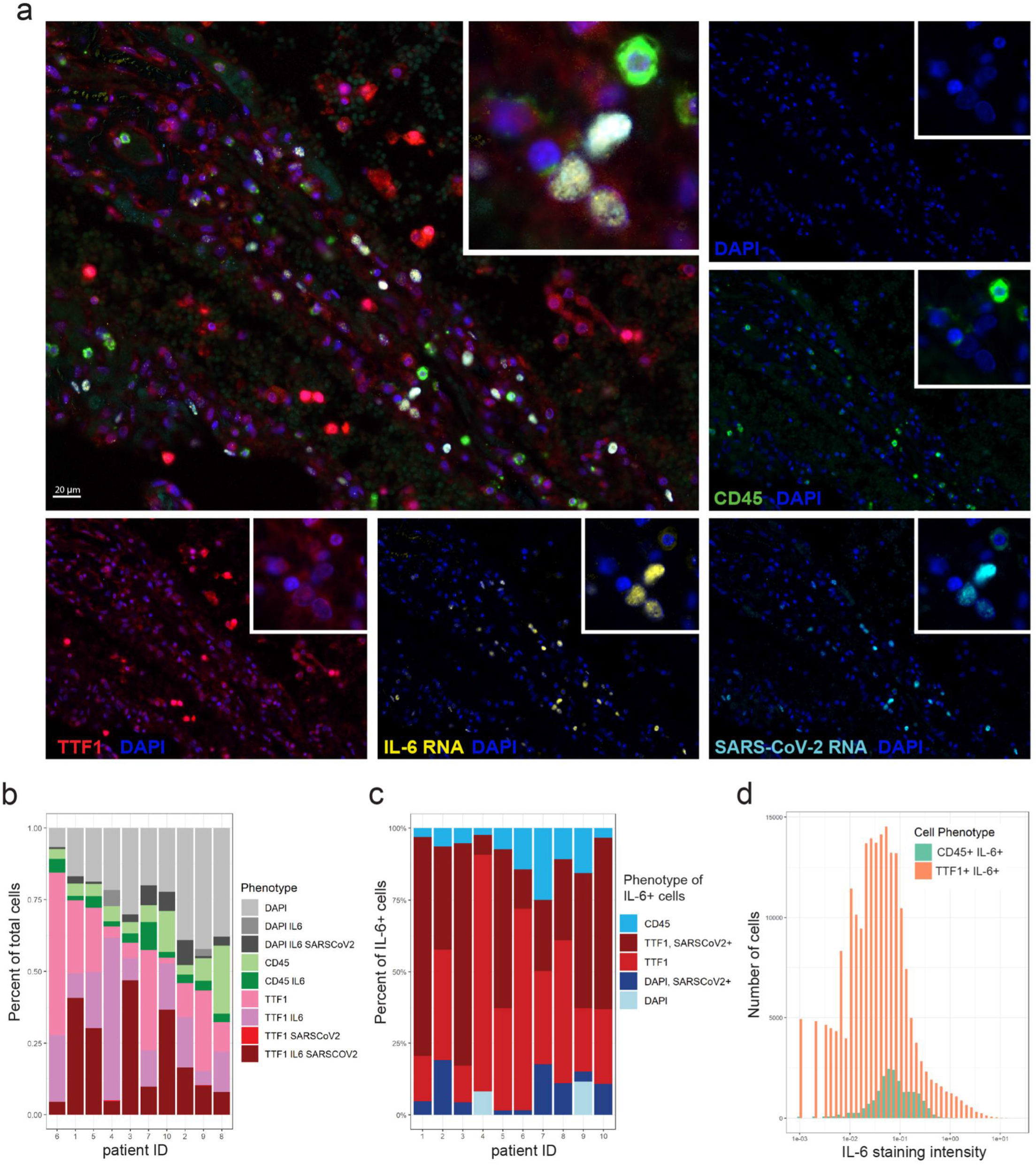
Lung epithelial cells predominantly express IL-6 in lung autopsy tissue in fatal COVID-19. Autopsy lung sections from 10 fatal COVID-19 cases were simultaneously stained for SARS-CoV-2 RNA, IL-6 mRNA, TTF1+ pneumocytes, and CD45+ leukocytes using RNA-ISH combined with multispectral immunofluorescence staining for protein. (a) Representative staining for TTF1 (red), CD45 (green), IL-6 RNA (yellow), SARS-CoV-2 RNA (light blue), and nuclear DAPI counterstain (blue); each stain shown separately and merged. Multispectral images were acquired at 40x magnification. Overlaying high-power images showing SARS-CoV-2 infected TTF1+ pneumocytes expressing high levels of IL-6. (b) Bar plots showing the phenotype composition of cell populations in each autopsy lung specimen. (c) Bar plots showing the phenotype composition of IL-6+ cells in each autopsy lung specimen. (d) Histogram displaying the frequency distribution of mean staining intensity for IL-6 between TTF1+IL-6+ cells (red) versus CD45+ IL-6+ cells (aqua). Cumulative data from all patients shown.

## Discussion

Here we show that IL-6 and CCL2 are major factors that discriminate severe infection from mild or moderate disease. IL-6 is known to be produced by innate immune cells such as macrophages or dendritic cells, and by non-immune cells such as epithelial cells or fibroblasts. In allergic asthma^44,45^, SARS-CoV-1^40^, influenza^41^, and pneumovirus infection models^42^, IL-6 has been shown to be produced by macrophages and other myeloid cells, whereas IL-6 can be produced by cultured nasal epithelial cells infected with RSV^46,47^. In mouse models of CAR-T cell cytokine release syndrome, macrophages and monocytes are the predominant source of IL-6^38,39^, while vascular endothelial cells have also been shown to produce IL-6 in CRS autopsy specimens^48^. Our results from human autopsy specimens unexpectedly show that the predominant source of IL-6 and CCL2 *in vivo* during SARS-CoV-2 infection is from infected epithelial cells. Our data are consistent with scRNA-seq studies of PBMCs from COVID-19 patients that showed a discrepancy between serum cytokine measurements and the cytokine transcripts of CCL2 and IL-6 among PBMCs^49–53^. Large numbers of epithelial pneumocytes co-stained with IL-6 or CCL2 and SARS-CoV-2 RNA probes, implicating direct cytokine induction by the virus. When considering potential mechanisms of cytokine production, it has been demonstrated that SARS-CoV-2 induces Nuclear Factor kappa B (NF-kB) upregulation and IL-6 production in cultured lung alveolar and epithelial cells^54,55^. CCL2 and other inflammatory mediators are also induced via the NF-kB pathway^56^.

In mouse models of coronavirus infections, sustained CCL2 expression enhanced the lethality of disease, and promoted immunopathology with a destructive monocyte/macrophage response and ineffective virus clearance^57^. The effect of excess CCL2 in human SARS-CoV-2 has not yet been elucidated. CCL2 may recruit wound-healing M2 macrophages, which can facilitate lung tissue repair by stimulating type 2 pneumocyte expansion^58^, thereby triggering a process capable of enhancing virus propagation^59^. The anti-IL-6R antibody tocilizumab improves survival in critically ill patients^9^, implying that excessive IL-6 is detrimental to the host. Elevated levels of IL-6 in cancer models have been mechanistically linked to decreased DC survival and activation, and consequently impaired CD8^+^ T cell priming^60^. As such, elevated IL-6 expression in the lung during SARS-CoV-2 infection might impair DC function within the infected lung. Thus, we speculate excess IL-6 and CCL2 may favor the virus by promoting a local defect in DC priming of T cells and impaired reactivation of virus-specific T cells locally within the lung and/or by supporting the survival and expansion of infected lung epithelial cells via recruitment of M2 wound-healing macrophages.

Corticosteroids, which improve survival for COVID-19 patients requiring supplemental oxygen^5^, exert their anti-inflammatory effects through NF-KB inhibition and other pathways^61^. It has been assumed that steroids are acting on immune cells, but they could also be inhibiting NF-kB in infected epithelial cells or other host cells. Together, these observations suggest a model whereby SARS-CoV-2 induces NF-kB, leading to an increase in IL-6 and CCL2 production in type 2 pneumocytes, creating favorable conditions for viral persistence, alveolar damage, and ultimately respiratory failure. Additional studies are needed to determine the impact of lung-derived IL-6 and CCL2 on immune clearance of SARS-CoV-2.

Robust adaptive immune responses were seen in patients with mild and moderate disease and were even higher in patients with severe disease, arguing that the lack of a protective immune response did not cause severe disease. CD38^+^HLA-DR^+^ CD4^+^ and CD8^+^ T cells were polyfunctional, and patients with a higher disease severity had more cytokine producing CD4^+^ T cells. These cytokines were being produced by PD-1^+^ cells, indicating that PD-1 in this context is a marker of activation, not exhaustion. This is consistent with other recent work showing that tetramer^+^ PD-1^+^, SARS-CoV-2-specific CD8^+^ T cells produce cytokines^62^. In patients with severe disease, markers of T cell activation such as sCD25/IL-2R remain high at late time points, suggesting an ongoing immune response against the virus. In some severe patients who ultimately die from SARS-CoV-2, persistent viral RNA has been demonstrated in longitudinal saliva samples from the Iwasaki group^63^, as well as in our autopsy lung samples. Increased antigen load and duration of antigenic exposure leads to increased T and B cell expansion and differentiation in other experimental models^64^. While we cannot rule out that the increased adaptive immune response causes immunopathology, the increase in regulatory modulators such as IL-10 and Tregs suggests that the immune system is appropriately executing negative feedback pathways. Our data suggests that the immune response to SARS-CoV-2 is a functional and proportional response to infection, and infected pneumocytes are the major source of IL-6 and CCL2, thus revising the paradigm of how we understand the pathogenesis of severe SARS-CoV-2 infection.

## Data Availability

All data produced in the present study are available upon reasonable request to the authors

---

**Supplementary Table 1:** Patients excluded from immune analysis.

|  |
| --- |
| **Reasons for exclusion of patients from immune analyses:** |
| Trauma/surgery during admission for SARS-CoV-2 |
| Systemic bacterial infection or bacteremia (or high clinical suspicion thereof) |
| Active malignancy |
| Biologic / Immune Suppressant use within the last 30 days $\geq 7.5$ mg of prednisone (or within last 6 months for long-acting monoclonal antibodies). Samples prior to receipt of steroids were eligible for inclusion. |
| Chemotherapy or Immunotherapy within the last 6 months |
| Sickle Cell Disease with crisis within the last 30 days |
| Solid organ transplant |
| Concurrent pneumothorax |
| Saddle pulmonary embolism with hemodynamic compromise |
| Cardiogenic shock in a patient with end stage heart failure; out of hospital cardiac arrest |
| Delivered of a pregnancy during admission for SARS-CoV-2 |

**Supplementary Table 2:**
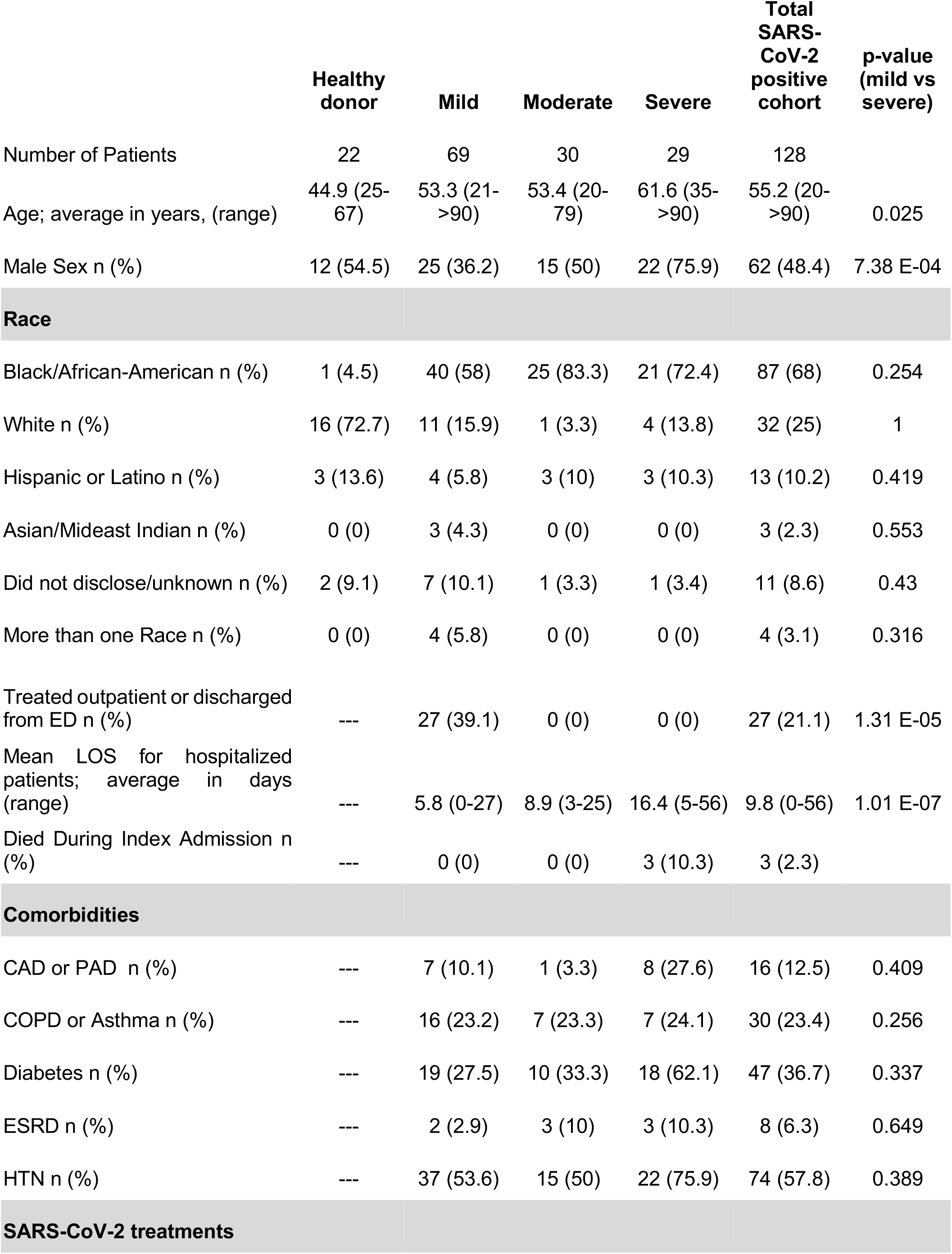
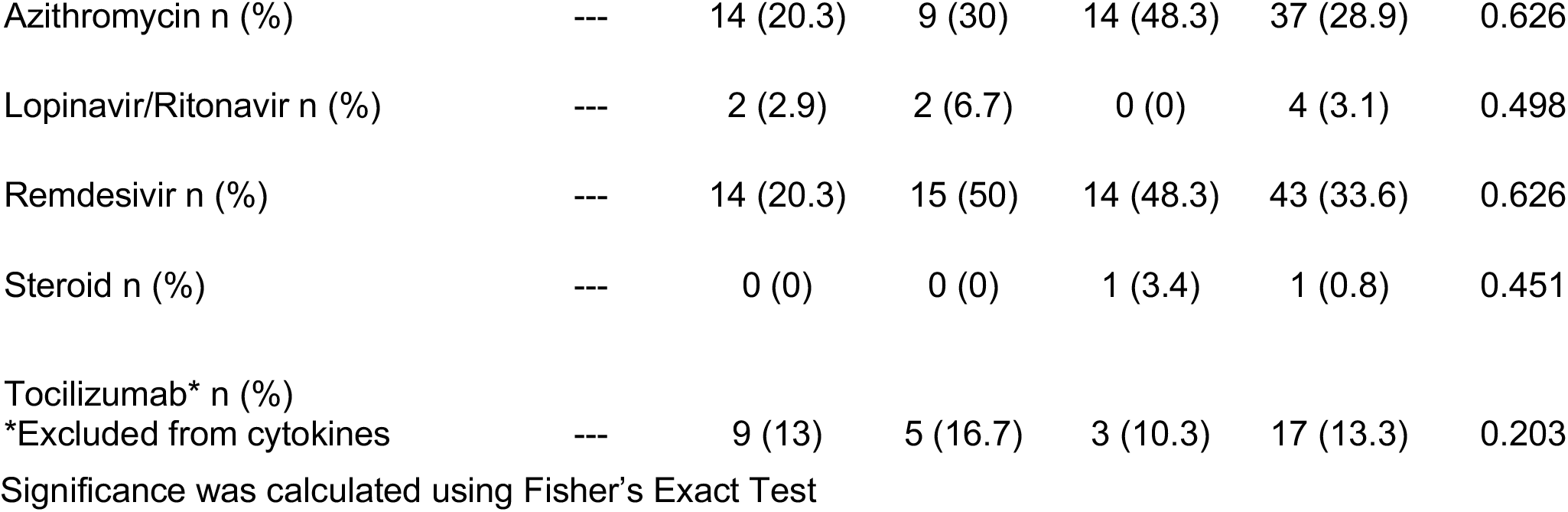
Demographic characteristics of patient cohorts.

**Supplementary Table 3.** Demographic characteristics of COVID-19 autopsy cases. Abbreviations used: AAA - Abdominal aortic aneurysm; CABG - coronary artery bypass graft; CAD - coronary artery disease; CKD - chronic kidney disease; CVA - cerebrovascular accident; DM - diabetes mellitus; HTN - hypertension; IDDM - insulin dependent diabetes mellitus; pHTN - pulmonary hypertension; OA - osteoarthritis; SLE - systemic lupus erythematosus.

| Patient ID | Sex | Age<br>(in decades for<br>anonymity) | Race/<br>ethnicity | Days<br>hospitalized<br>prior to<br>death |
| --- | --- | --- | --- | --- |
| 1 | M | 30-40 | AA | 9 |
| 2 | F | 50-60 | Hispanic | 12 |
| 3 | F | 80-90 | AA | 13 |
| 4 | F | 60-70 | AA | 4 |
| 5 | F | 80-90 | AA | 4 |
| 6 | M | 80-90 | AA | 1 |
| 7 | M | 40-50 | AA | 21 |
| 8 | F | 70-80 | AA | 15 |
| 9 | F | 80-90 | Caucasian | 1 |
| 10 | F | 70-80 | AA | 4 |

## Acknowledgements

We would like to thank Marcellus Johnson, Melanie Veron, and Lauren Wall for clinical research support, Hongyuan Jiao, PhD, Glee Guilan Li, and Shuhan Yu, MD for biobank sample processing, as well as Rajlakshmi Krishnamurthy MD and Geneatra Green for operational support. Thanks to Laura Johnson from the flow cytometry core, Melvin Lye from Curiox, and Chris Fleming from Cytek for assistance with assay design. The HIM and flow cytometry cores are supported by the University of Chicago Cancer Center Support Grant P30CA014599, and the Center for Research Informatics and Redcap are supported by the UL1TR000430 from NCATS/NIH. SJR, JY and CSC are supported by the T32 CA009566. JAT is supported by 5K12CA139160-09 from NIH. RR was funded by the German Research Foundation (DFG RE 4468/1-1). MLM received funding from NIAID Collaborative Influenza Vaccine Innovation Centers (CIVIC) contract 75N93019C00051. YL and LSC are supported by R01GM108711.

## Author Contributions

SJR, JAT, ARP, TFG conceived and designed the study. SJR, JAT, ARP, AP, LK, GWS, CSC, MK, SS, and MLM consented and recruited patients. YZ processed and stored patient samples and isolated RNA. SJR, JAT, ARP, JF, KC performed flow cytometric assays; AC and JCB performed ELISAs; AC, JAT, BAF, YZ performed cytokine assays; JAT, KH, RR performed microscopy, MFA and SO performed ddPCR. YSJR, JAT, ARP, JY, JF, AC, EFH, KC, YL, JCB, MFA, SO and BAF analyzed data. JY, EFH, YL, and BAF performed bioinformatic analysis, did statistics, and graphed data. ML, KB, K-TJY, JM provided materials and samples. RS, ST, EI, JS, LC, TFG provided personnel, supervision, and insights. SJR, JAT, ARP, and TFG drafted the manuscript; all authors helped to edit the manuscript.

Correspondence and requests for material or data should be directed to TFG.

## Methods

### Patients, sample collection

The COVID-19 biobank study was approved by the University of Chicago Institutional Review Board (IRB 20-0520), and all procedures were performed in accordance with the ethical standards set forth in the 1964 Helsinki Declaration. Informed consent was obtained using paper consent, or a Redcap electronic consent form (when possible) to minimize risk of infection. Patients could choose to donate fresh research blood samples and/or allow leftover material from clinical testing (BMP and nasopharyngeal swabs) to be used for research. Research samples were used for flow cytometry, ELISPOTs, and luminex analysis; ELISAs used both research plasma samples and leftover serum from clinical BMPs. Viral load was measured on leftover viral transport media from clinical nasopharyngeal swabs. Blood was collected from inpatients every 24-72 hours during the first week of their hospitalization, and 1-2 times a week for the remainder of their hospitalization. Blood was collected up to once a week from outpatients during regularly scheduled appointments. Serum from 5 healthy donors was purchased from Cellular Technology Limited; the remainder of the healthy donor samples were obtained from the COVID-19 biobank.

For serum, fresh blood was collected into a preservative-free vacutainer tube and allowed to clot for at least 30 minutes at room temperature. Leftover plasma samples collected in heparinized tubes were also obtained from the clinical chemistry lab. Tubes were centrifuged for 20 minutes at 1300 x g at room temperature, and the yellow serum/plasma layers were collected and stored at -80 degC until analysis.

For PBMCs, fresh blood was collected into heparinized vacutainer tubes and separated using LeucoSep (Greiner Bio-One) tubes. Cells were washed twice with PBS, counted and resuspended in freezing media containing 90% FBS and 10% DMSO at 5 – 20×10^6/mL. Cells were frozen in liquid nitrogen until further analysis.

### Timepoints used

For analyses divided into early (D0-9) and late (D10-30) phases, the maximal cytokine measurement per patient and the mean of any available flow cytometry measurements per patient in each time period were used. For kinetic analysis of cytokines or antibodies versus DPSO, all available measurements between D1-30 per patient were used. For cytokines versus total non-mild days, the maximal cytokine value between D1-30 was used. Total non-mild days was calculated as the number of hospitalized days with an average S/F ratio of < 315. For maximal antibody titers, the highest antibody titer measurement between D10-30 per patient were used. Maximum viral load per patient was used. ICS samples were from D11-17 DPSO.

### Cytokine measurements by Luminex

Human XL Cytokine Luminex Performance Panel Premixed Kit (CCL2/JE/MCP-1, CCL3/MIP-1 alpha, CCL4/MIP-1 beta, CCL5/RANTES, CCL19/MIP-3 beta, CCL20/MIP-3 alpha, CX3CL1/Fractalkine, CXCL1/GRO alpha/KC/CINC-1, CXCL2/GRO beta/MIP-2/CINC-3, CXCL10/IP-10/CRG-2, EGF, G-CSF, GM-CSF, Granzyme B, IFN-alpha, IFN-gamma, IL-1 alpha/IL-1F1, IL-1 beta/IL-1F2, IL-1ra/IL-1F3, IL-2, IL-6, IL-7, IL-10, IL-13, IL-15, IL-33, TGF-alpha, TNF-alpha, TRAIL/TNFSF10, VEGF) and human 6plex luminex multiple kits (angiopoietin-2, CD25/IL-2R alpha, gp130, IL-6R alpha, VEGF-c) were purchased from Bio-Techne, and performed according to manufacturer’s instructions. Samples were run in duplicate. Any analytes with a bead count < 32 or CV > 20% between the duplicates was considered to fail QC for that patient and excluded from further analysis.

### Anti-SARS-CoV-2 antibody ELISA

The SARS-CoV-2 full-length Spike and Receptor Binding Domain (RBD) protein expression constructs were obtained from Florian Krammer and Patrick Wilson^65^, and used to generate recombinant protein for an enzyme-linked immunosorbent assay (ELISA) adapted from established protocols^66^. Recombinant proteins were produced using a Chinese hamster ovary (CHO) cell line expression system and purified using metal-chelate affinity chromatography. Protein integrity was confirmed via SDS-PAGE gel. Overnight, 96-well ELISA plates (Nunc MaxiSorp high protein-binding capacity plate; ThermoFisher) were coated at 4°C with 2 mg/mL of Spike or RBD protein suspended in Phosphate Buffered Saline (PBS) pH 7.4. Plates were blocked with 3% milk powder in PBS containing 0.1% Tween-20 for 1 hour at room temperature. Serum and plasma samples were heated at 56°C for 30 minutes to inactivate virus prior to use. Serial 1:3 dilutions of the samples were prepared in 1% milk in 0.1% PBS-Tween 20, and incubated in duplicate with the blocked plate for 2 hours at room temperature. After 3 washes in 0.05% PBS-Tween 20, an HRP-conjugated secondary antibody specific for either total Ig (goat anti-human immunoglobulin Ig, SouthernBiotech), IgM (anti-human IgM, μ-chain specific, Millipore Sigma), or IgG (Goat anti-human IgG (H+L), Jackson ImmunoResearch) were diluted in 1% milk in 0.1% PBS-Tween-20, and added at 1:8000 for 1 hour at room temperature. Plates were washed 3 times with 0.1% PBS-Tween 20 before being developed with 3, 3’, 5, 5’-tetramethylbenzidine (TMB) substrate kit (ThermoFisher) at room temperature. The reaction was stopped after 15 minutes with 2M sulfuric acid. The optical density (OD) was read at 450 nm using a Synergy H4 plate reader (BioTek). The OD values for each sample were background subtracted. A positive control standard was prepared from plasma samples pooled from 6 COVID-19-infected patients, while plasma from an uninfected individual was used as a negative control standard. To account for variability between plates, the OD values were divided by the OD from the negative control from each plate, run at a 1:50 dilution. To quantify the amount of anti-Spike Ig and anti-RBD Ig in the sample, end-point titers were calculated as the linear interpolation of the inverse dilution at which the normalized OD value crossed a threshold of 1, which was the maximum OD measured for the negative control.

### Flow cytometry

Frozen PBMCs were thawed into 10 mL RPMI with 10% FCS, 1 mM EDTA + DNAse I, washed, and plated on 96 well laminar wash plates (Curiox). Subsequent washes were done using a laminar flow plate washer (Curiox) in a BSL-2 hood, and all staining was done at room temperature. Cells were allowed to settle for 40 minutes, washed, and stained with Live/Dead Blue (Invitrogen) in PBS for 20 minutes. Cells were washed and incubated with monocyte blocker (Biolegend) and CCR7 BV421 (G043H7, Biolegend, 5 uL) for 10 minutes. Brilliant stain buffer plus (BD), CXCR3 PECy7 (G025H7, Biolegend, 5 uL), CXCR5 BV750 (RF8B2, BD, 1.2 uL), CCR6 BV711 (G034E3, Biolegend, 1.2 uL), and TCRgd PCPCeF710 (B1.1, Thermo, 5 uL) were added for 10 minutes, and the remainder of extracellular antibodies (CD101 BUV563 (V7.1, BD, 5 uL), CD11b PCPCy5.5 (ICRF44, Biolegend, 0.6 uL), CD11c BUV661 (B-ly6, BD, 3.5 uL), CD123 Super Bright 436 (6H6, Thermo, 3.5 uL), CD127 APC-R700 (HIL-7R-M21, BD, 6 uL), CD14 SparkBlue550 (63D3, Biolegend, 2.5 uL), CD141 BB515 (1A4, BD, 2.5 uL), CD15 Pacific Orange (VIMC6, Thermo, 5 uL), CD16 BUV496 (3G8, BD, 0.6 uL), CD161 eFluor450 (HP-3G10, Thermo, 5 uL), CD19 SparkNIR685 (HIB19, Biolegend, 2 uL), CD1c AF647 (L161, Biolegend, 5 uL), CD25 PE (BC96, Biolegend, 10 uL), CD27 APC (M-T271, Biolegend, 5 uL), CD28 BV650 (CD28.2, Biolegend, 2.5 uL), CD3 BV510 (OKT3, Biolegend, 5 uL), CD38 APC-Fire810 (HIT2, Cytek/Biolegend, 1 uL), CD4 CFluor568 (SK3, Cytek, 1.2 uL), CD45 PerCP (2D1, Biolegend, 1.2 uL), CD45RA BUV395 (5H9, BD, 0.3 uL), CD45RO BV605 (UCHL1, Biolegend, 5 uL), CD56 BUV737 (NCAM16.2, BD, 3.5 uL), CD57 FITC (HNK-1, Biolegend, 1.2 uL), CD8 BUV805 (SK1, BD, 1.2 uL), CD86 BUV615 (BU63, BD, 5 uL), HLA-DR APCF750 (L243, Biolegend, 2.5 uL), CD95 PECy5 (DX2, Biolegend, 0.6 uL), IgD BV480 (IA6-2, BD, 0.6 uL), IgM BV570 (MHM-88, Biolegend, 2.5 uL), PD-1 BV785 (EH12.2H7, Biolegend, 5 uL), TIM-3 PEDz594 (F38-2E2, Biolegend, 5 uL)) were added for 45 minutes. Cells were washed and resuspended in 2% paraformaldehyde for 30 minutes, and then washed and run on the Cytek Aurora spectral flow cytometer. Data analyzed using FlowJo.

### Intracellular cytokine staining

Frozen PBMCs were thawed into T cell media (RPMI with 10% FBS, 2 mM L-glutamine, 1 mM sodium pyruvate, 50 uM 2-BME, 100 U/mL penicillin, 100 mg/mL streptomycin) washed, and allowed to rest overnight. PepTivator SARS-CoV-2 peptide pools consisting of 15-mer sequences with 11 amino acids overlap against the immunodominant sequence of the surface glycoprotein (“ S”), and complete sequences of the membrane glycoprotein (“ M”) and nucleocapsid phosphoprotein (“ N”) (all from Miltenyi Biotec) were combined and used at 1 ug/mL, along with 0.5 ug anti-CD28/CD49D antibodies (clone L293/L25, BD biosciences). The PepTivator SARS-CoV-2 peptide pools stimulate both CD8^+^ and CD4^+^ T cells. For αCD3/CD28/CD49d, plates were coated overnight at 4 degC with 10 ug/mL anti-CD3 (clone SK7) in PBS, and 0.5 ug anti-CD28/CD49d was added into the media with the PBMCs. 0.1 ug/mL phorbol myristate acetate (PMA) and 1 ug/mL ionomycin were to the appropriate wells with PBMCs. The unstimulated condition was also treated with 0.5 ug anti-CD28/CD49d. All conditions were incubated for 9 hours in the presence of Golgiplug/Golgistop at manufacturer’s recommended concentration (BD Biosciences). After stimulation, 2 mM EDTA was added to the anti-CD3/CD28/CD49d wells and incubated for 15 minutes. Cells were then transferred to a V-bottom 96 well plate for staining. Cells were stained with live/dead blue (Invitrogen) in PBS for 15 minutes prior to adding monocyte blocker and CCR7 BV421 (G043H7, Biolegend, 5 uL) for 10 minutes. The remainder of the extracellular antibodies (TCRgd PCPCeF710 (B1.1, Thermo, 5 uL), CD3 BV510 (OKT3, Biolegend, 5 uL), PD-1 BV785 (EH12.2H7, Biolegend, 5 uL), CD56 BUV737 (NCAM16.2, BD, 3.5 uL), HLA-DR APCF750 (L243, Biolegend, 2.5 uL), CD20 Pacific orange (HI47, Thermo, 5 uL), CD4 CFluor568 (SK3, Cytek, 1.2 uL), CD45 PerCP (2D1, Biolegend, 1.2 uL), CD19 SparkNIR685 (HIB19, Biolegend, 2 uL), CD8 BUV805 (SK1, BD, 1.2 uL), CD38 APC-Fire810 (HIT2, Cytek/Biolegend, 1 uL), CD16 BUV496 (3G8, BD, 0.6 uL), CD45RA BUV395 (5H9, BD, 0.3 uL)) and brilliant stain buffer plus (BD) in a total volume of 100 uL were added for 30 minutes, and cells were washed once. Cells were resuspended in the fixation/permeabilization solution from eBioscience’s FoxP3 / transcription factor staining buffer for 30 minutes, washed, and incubated with intracellular antibodies in 100 uL (IFNy AF488 (4S.B3, Biolegend, 5 uL), Granzyme AF532 (N4TL33, Thermo, 5 uL), IL-10 APC (JES3-19F1, Biolegend, 5 uL), TNFa BV605 (MAb11, Biolegend, 5 uL), IL-2 BV650 (MQ1-17H12, Biolegend, 5 uL), Ki67 Pacific blue (Ki-67, Biolegend, 5 uL), IL-17A PE (BL168, Biolegend, 5 uL), IL-4 PEDz594 (MP4-25D2, Biolegend, 5 uL), IL-6 PECy7 (MQ2-13A5, Biolegend, 5 uL), Perforin BV711 (dG9, Biolegend, 5 uL)) for 60 minutes in permeabilization buffer. Cells were washed 2x with permeabilization buffer prior to being resuspended in FACS buffer and run on the Cytek Aurora. Data analyzed using FlowJo.

### ELISPOT

PBMCs were thawed and washed with 10 mL RPMI prior to being plated in precoated human IFN-γ ELISPOT plates (ImmunoSpot). Control MHC I and II peptide pools against common viral antigens were used as positive controls (MHC I: 2 ug/mL of CEF peptide pool plus against CMV, EBV, and influenza; MHC II: 50 ug/mL of CPI pool against CMV, influenza and parainfluenza; both from ImmunoSpot). Wells with 0.1 ug/mL PMA and 1 ug/mL ionomycin were used as an additional positive control. PBMCs in separate wells were stimulated with peptides from either the spike protein (“ S”), membrane glycoprotein (“ M”) or nucleocapsid phosphoprotein (“ N”) (all from Miltenyi Biotec) at 1 ug/mL. SIYRYYGL (SIY peptide) was used as a negative (irrelevant) peptide control in each experiment with ≤1 spot seen in each irrelevant well. The total number of PBMCs recovered after thawing was divided into 12 wells, with PMA + Ionomycin controls plated at 10% of the cell density as other wells. Average cell number per experimental well was 218,000, with a range of 50,000 - 750,000 cells for experimental wells. Cells were incubated at 37°C with 5% CO2 with activating stimuli for 18 hours in CTL-Test Medium (ImmunoSpot) with 1% L-glutamine (Gibco) prior to developing plates per manufacturer’s recommended procedure. Plates were scanned using ImmunoSpot analyzer and spots were counted using ImmunoSpot 7.0.17.0. Spots were normalized by dividing by number of cells plated per well * 100,000 to report spots per 100,000 cells. The sum of the response in the S + M + N wells per 100,000 cells was reported as IFN-γ spots / 100,000 cells.

### Viral load

Leftover viral transport media from clinical nasopharyngeal swabs was stored at -80°C until analysis. RNA was extracted using the Qiagen viral RNA mini kit following the manufacturer’s instructions, with RNA eluted in 60 μL of AVE buffer. Digital droplet PCR (ddPCR) was performed as previously described^67^. Briefly, a 20 uL reaction was performed with 2 uL N1/N2/RNaseP probe primer sets (IDT #10006770), 5 uL one-step ddPCR supermix, 2 uL reverse transcriptase, 1 uL 300 mM DTT, and 6 uL sample RNA. Amplification was performed under the following conditions: 25°C for 2 min, 50°C for 60 min, 95°C for 10 min, 45 cycles of [95°C for 30 seconds, 55°C for 1 min, then 98°C for 10 min], followed by infinite hold at 4°C. Ramping speed was 2.5°C/s. Fluorescence was read using a QX200 droplet reader (Bio-Rad) and analyzed with Quantasoft software. Threshold of positivity defined in the manufacturer’s FDA emergency use authorization approval was used (>0.1 copy number / μL for either N1 or N2 and more than 2 positive droplets per reaction). Viral load is reported as the average of the N1 and N2 copy number / μL.

### Multiplexed staining combining RNA-ISH and immunofluorescence staining for protein

Simultaneous detection of RNA and protein antigens was performed by combining RNA *in situ* hybridization (ISH) using the RNAScope® Multiplex Fluorescent Reagent Kit v2 assay together with antibody-based immunofluorescence staining, according to the manufacturer’s integrated co-detection protocol (Advanced Cell Diagnostics, ACD). Briefly, formalin-fixed paraffin-embedded (FFPE) lung tissue sections were baked for 30 min at 60° C, deparaffinized by submerging in xylenes for 5 mins twice. The sections were rehydrated in 100% ethanol for 1 min twice, air dried, treated with RNAScope® hydrogen peroxide for 10 min, and rinsed with distilled water. Target retrieval was performed in a TintoRetriever Pressure cooker (Bio SB) using 1x Co-Detection Target Retrieval solution at 98-102° C for 15 min. Slides were rinsed in distilled water and 1x Phosphate-Buffered Saline Tween buffer (PBST). The tissue sections were blocked with Co-Detection antibody diluent (ACD) and incubated with anti-TTF1 antibody (SPT24, BioCare Medical) overnight at 4° C, washed in PBS-T buffer, incubated in 10% Neutral Buffered Formalin for 30 min at RT, and washed in PBS-T. Tissue sections were treated with RNAscope® Protease plus at 40° C for 30 min and rinsed in distilled water. *In situ* hybridization was performed according to the RNAScope® assay protocol. Briefly, sections were incubated with RNA probe mix and hybridized at 40° C for 2 hours. The following RNAscope® probes were used: V-nCoV2019-S-C1 (specific for SARS-CoV-2, S gene encoding the spike protein), Hs-IL-6-C4, Hs-CCL2-C2. Signal amplification was performed using the RNAScope Multiplex FL v2 AMP reagents, AMP1 (30 min, 40° C), AMP2 (30 min, 40° C), and AMP3 (15 min, 40° C), sequentially. Development of the horseradish peroxidase (HRP) signal was performed according to the manufacturer’s protocol. Fluorescent labeling of the IL-6 and CCL2 RNA probes was performed using OPAL 620 dye (Akoya Biosciences), while labeling of the SARS-CoV-2 RNA probe was performed using Opal 540 dye. The TTF1 primary antibody was detected with HRP-conjugated secondary antibody (Opal Polymer HRP Ms + Rb, Perkin Elmer) and Opal 690 dye. Subsequent staining on the same sections was performed with an antibody against CD45 (Leukocyte Common Antigen Cocktail: PD7/26/16 and 2B11, BioCare Medical) and detected with HRP-conjugated secondary antibody and Opal 520 dye. After all targets were labeled, the sections were incubated with DAPI solution for 5 min at room temperature and mounted in ProLong Diamond Antifade Mountant (Invitrogen). Co-staining for IL-6 RNA and macrophages was performed using RNAscope® probe Hs-IL-6, which was fluorescently labeled with OPAL 690 dye, and an antibody against either CD68 (Clone KP1, BioCare Medical; diluted in Da Vinci Green diluent) or CD163 (Clone 10D6, BioCare Medical; diluted in Renoir Red diluent) and detected with HRP-conjugated secondary antibody (Opal Polymer HRP MS+ RB) and OPAL 540 dye. Slides were scanned using the Vectra Polaris imaging platform and Phenochart software (PerkinElmer). For quantitative analysis, up to 200 representative fields of view for each tissue section were acquired at 40x magnification as multispectral images. Image analysis and cell phenotyping were performed using a supervised machine learning algorithm within the Inform 2.3 software (PerkinElmer), which assigned trained phenotypes and cartesian coordinates to cells.

### Clinical data warehouse

Clinical data was exported through a clinical data warehouse and was also abstracted by a clinical data manager into a standardized RedCap database. Patients with a history of active cancer, organ transplantation were excluded. Medications administered were searched from the clinical data warehouse to find and exclude patients receiving steroids, immunosuppressants, chemotherapy, or immunotherapy agents. Patients with positive blood cultures were identified and excluded. Patients noted to have alternative causes of cardiovascular shock or a pneumothorax were excluded as indicated in Table 1. Outpatients did not routinely have FiO2 documented, so outpatients were all categorized as mild with an S/F ratio set to 476 (equivalent to SpO2 100% on room air).

### Statistics and Data processing

Throughout the paper, boxplots show the medians (middle line) and the first and third quartiles (upper and lower bounds of the boxes). Significance of comparisons from boxplots were determined by two-sided Mann Whitney Wilcoxon test and significance is expressed as p-values, shown as asterisks (*, p ≤ 0.05; **, p ≤ 0.01; ***, p ≤ 0.001). All replicates are from distinct samples. Correlations between clinical and research parameters were analyzed using pairwise Spearman’s correlation coefficients and were visualized with R package corrplot^68^. Correlation was quantified by a color scale, the significance of the correlation was labeled with asterisks, and boxes with a thick black border represent a false-discovery rate (FDR) <0.05.

To determine which features were predictive of disease severity, the Random Forest R package randomForest^69^ was used and area under the ROC curve (AUC) calculated. A 3-fold cross-validation was used and the mean AUC on the test data set was presented. Models were trained for early phase (D0-D9), late phase (D10-D30), and all timepoints (D0-D30) separately. Correlation plots and the machine learning algorithm were processed using R studio (R 3.6.1).

Flow cytometry data were processed using FlowJo V10.7.1. Graphs were created and statistics performed using either GraphPad Prism v9.0.0 or R 4.0.3 (R Core Team, 2020). R packages used include Formula^70^, ggpmisc^71^, ggpubr^72^, ggsignif^73^, gridExtra^74^, Hmisc^75^, lubridate^76^, magrittr^77^, patchwork^78^, readxl^79^, reticulate^80^, rstatix^81^, scales^82^, stringr^83^, survival^84^, tidyverse^85^, writexl^86^, zoo^87^, and corrplot^68^.

## Data Availability

Raw .fcs files are available at (will be deposited prior to publication). The datasets generated during and/or analysed during the current study are available from the corresponding author on reasonable request.

## Code Availability

Full reproducible code for data processing is available at https://github.com/jovianyu/covid19biobank.

---

**Supplemental Fig 1:**
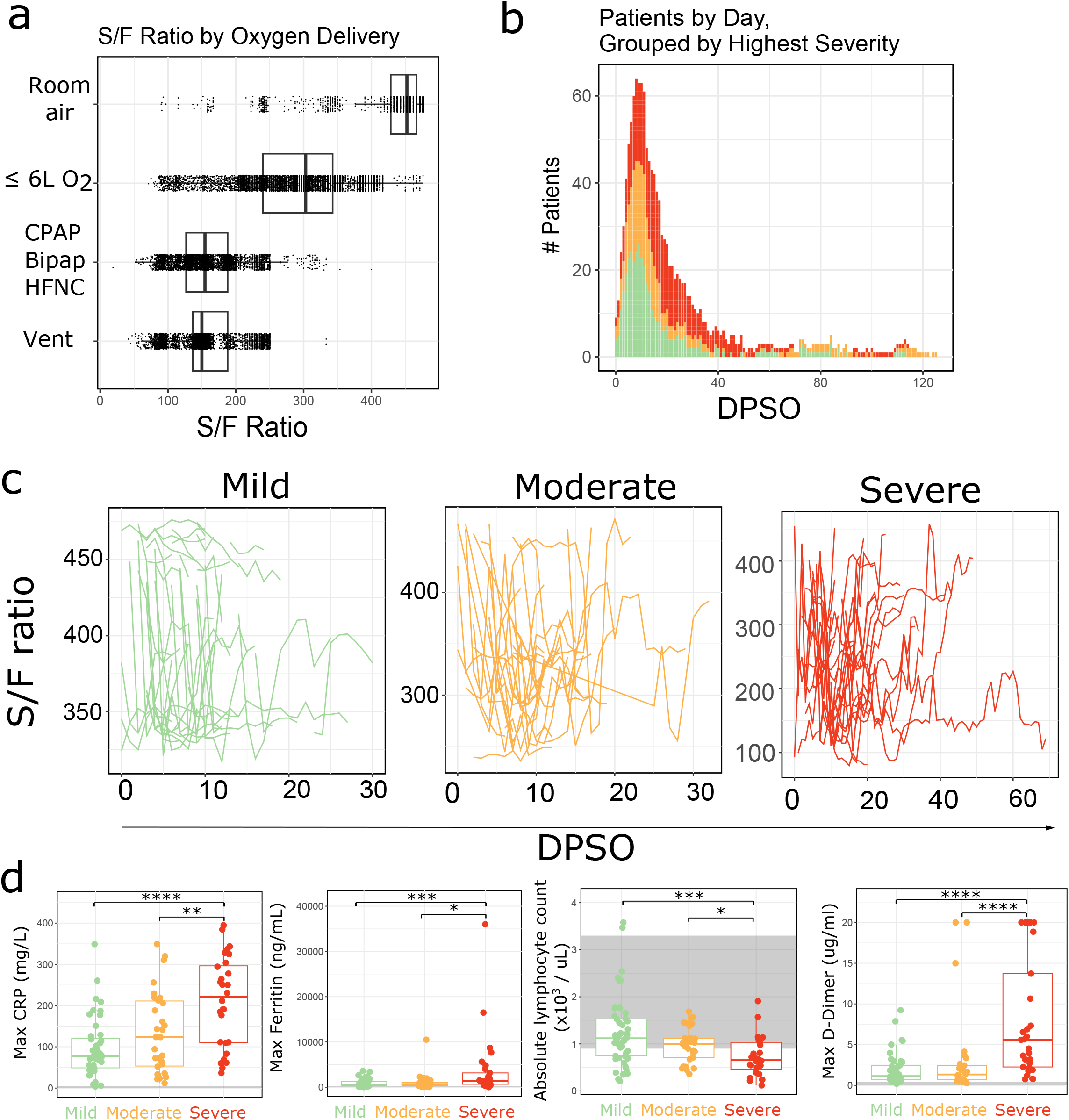
Clinical features of study cohort (a) The relationship between S/F ratio and oxygen delivery is depicted. All values during initial admission for all patients (n = 126) are included. Each dot represents an individual S/F ratio, and multiple ratios per patient per day are plotted. This includes individual values with the SpO2 < 88% which preceded an increase in the patient’s clinical oxygenation requirement. (b) Stacked histogram showing number of patients hospitalized versus days post symptom onset; each patient is represented multiple times based on duration of hospitalization. Disease severity is indicated by color (mild - green, moderate - orange, severe - red). (c) Mean daily S/F ratio is shown for mild, moderate and severe patients over time, with data from the same patients connected by lines. (d) Maximum CRP, Ferritin, D-Dimer and nadir of absolute lymphocyte count are shown for mild, moderate, and severe patients. Each dot represents the maximum or nadir value for each patient over the course of their initial hospitalization. The grey line indicates the maximum normal value. The grey block background indicates the normal reference range. Significance was determined by two-sided Mann Whitney Wilcoxon test and p-values are indicated by asterisks (*, p ≤ 0.05; **, p ≤ 0.01; ***, p ≤ 0.001, ****, p ≤0.0001).

**Supplemental Fig 2:**
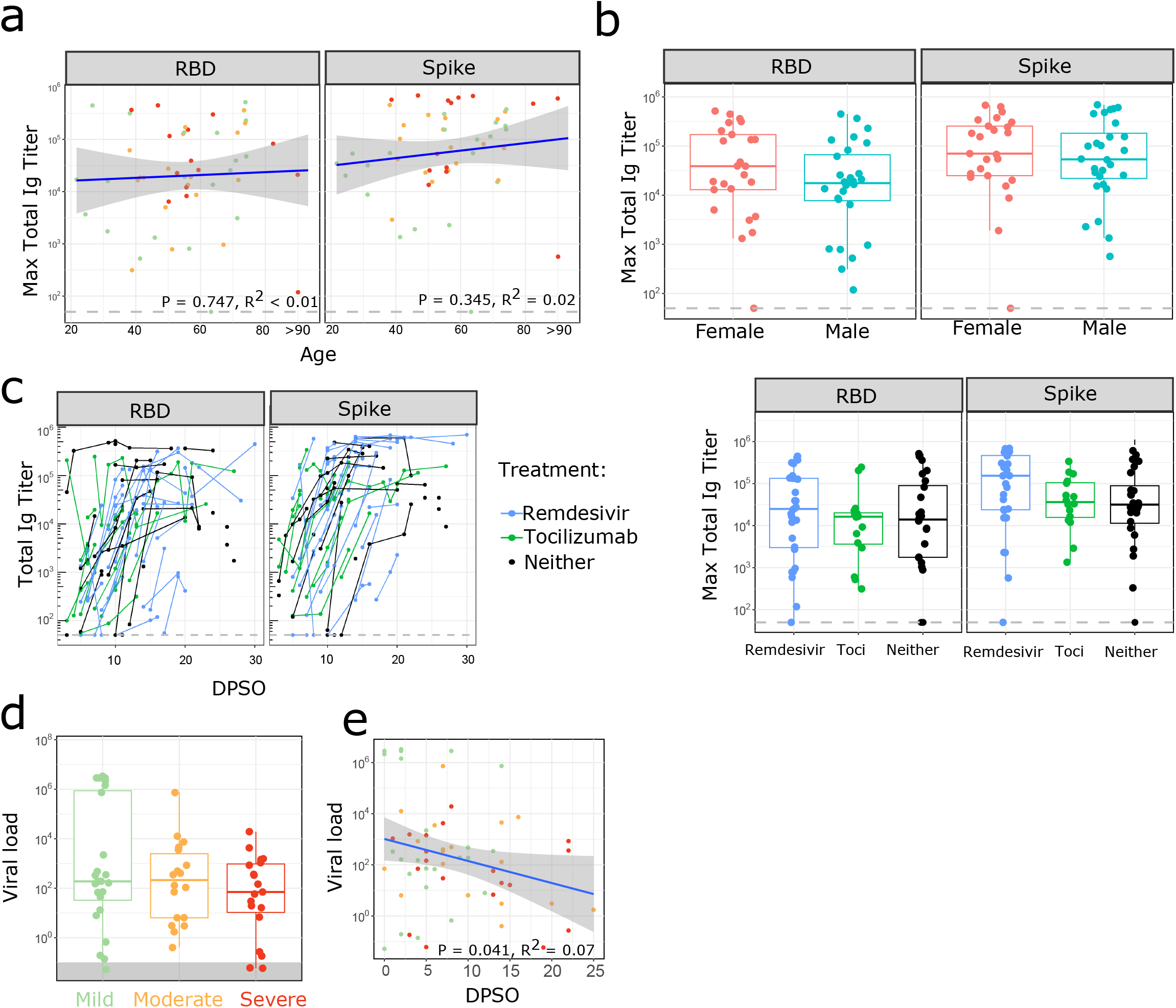
Antibody response does not differ based on age, gender, or treatment with remdesivir or tocilizumab (a) Maximum serum SARS-CoV-2 total Ig antibody levels (RBD and Spike) between D10-30 in PCR+ patients (n = 53) are shown by age. Disease severity indicated by color (green = mild, orange = moderate, red = severe). (b) Maximum serum SARS-CoV-2 total Ig antibody levels (RBD and Spike) between D10-30 in PCR+ patients are shown for females (F, n=25) and males (M, n=28). (c) Serum SARS-CoV-2 total Ig antibody levels (RBD and Spike) in PCR+ patients receiving remdesivir (blue, n=28), tocilizumab (green, n=16), or neither (black, n=24). Antibody levels over time shown in the left panel, and maximum titer per patient shown in the right panel. (d-e) Viral titers were obtained by ddPCR on leftover viral transport medium. Disease severity indicated by color (green = mild, n=24; orange = moderate, n=18; red = severe disease, n=19) patients. In d), the grey shaded area denotes negative results. (e) Linear correlation between day post symptom onset and viral load measured by ddPCR, disease severity is indicated by dot color. (a,e) Shaded areas represent 95% confidence interval. (b,c,d) Groups were compared with two-sided Mann Whitney Wilcoxon test and no significant differences were seen.

**Supplemental Fig 3:**
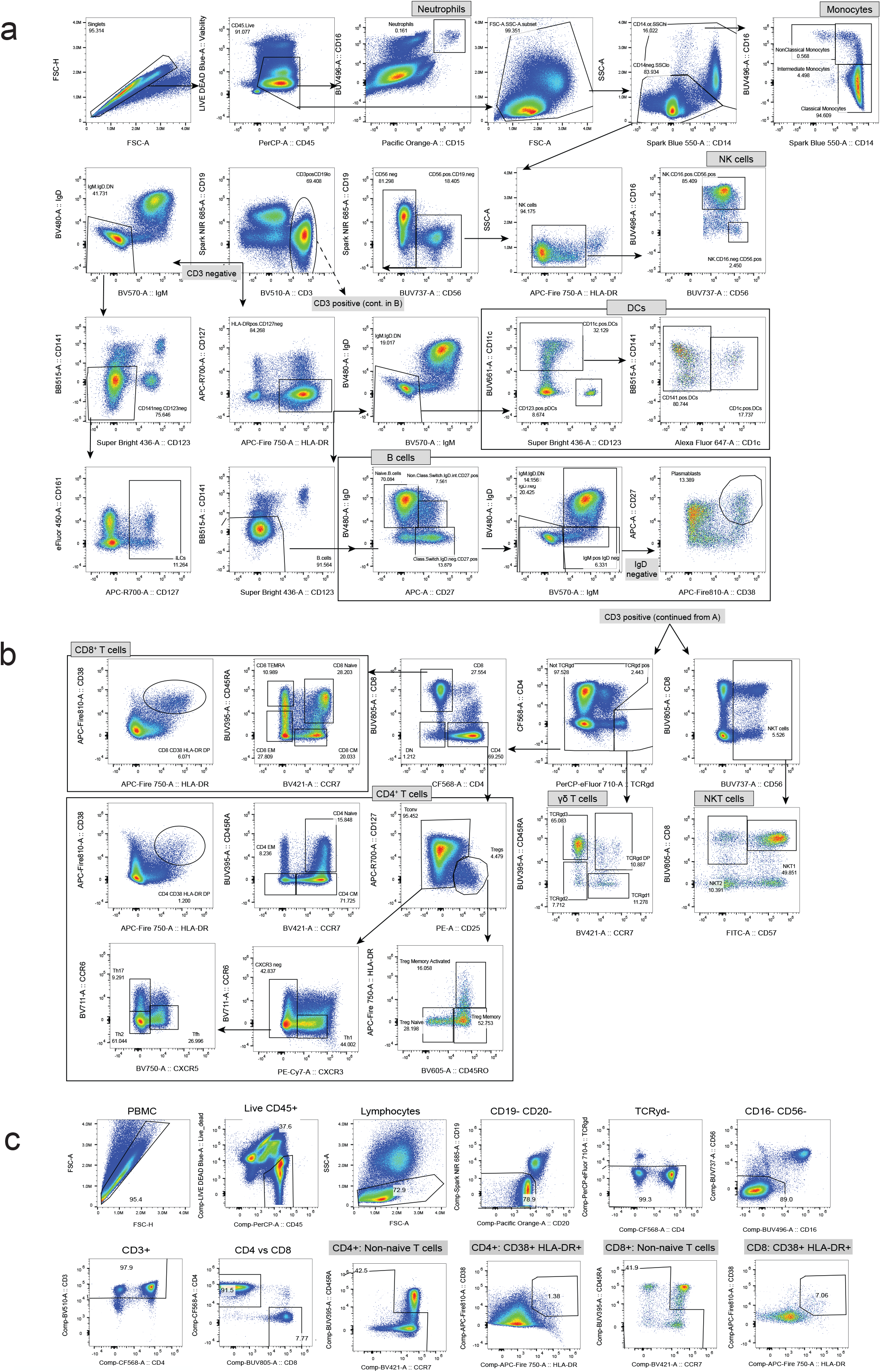
Flow cytometry gating strategy. (a) Gating for extracellular flow cytometry for monocytes, dendritic cells, B cells, and NK cells. (b) Gating for extracellular flow cytometry for T cells, γδ T cells, and NKT cells. (c) Flow cytometry gating for intracellular cytokine stimulation assays.

**Supplemental Fig 4:**
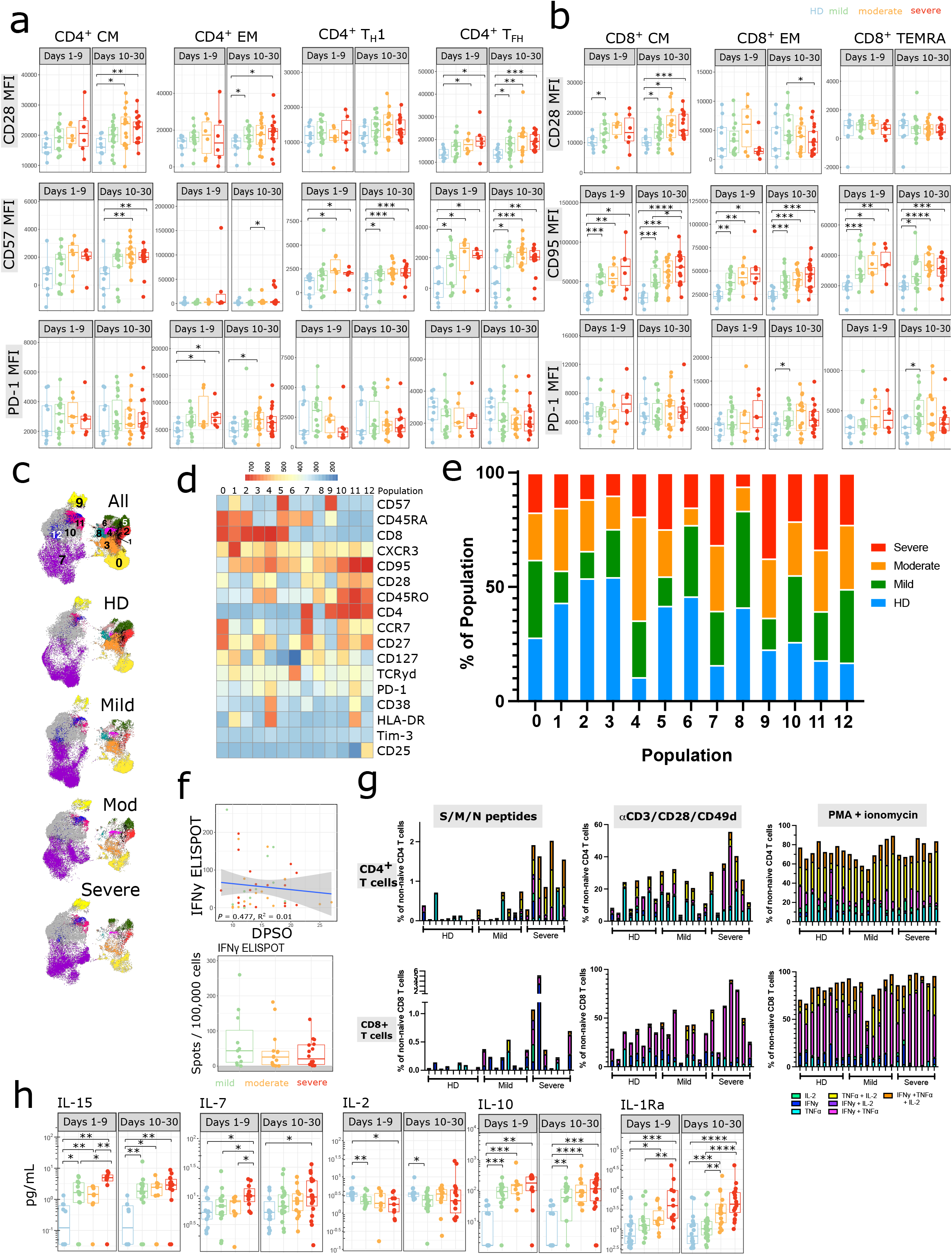
SARS-CoV-2 infection is associated with increased activation and functionality of CD4+ and CD8+ T cells. (a-b) Boxplots show MFI of indicated markers in each CD4+ (a) or CD8+ (b) T cell population. Where multiple timepoints within the early (D1-9) or late (D10-30) phase per patient were available, the mean was taken and each patient is represented by one dot per time phase. n = 9 for HD (blue), n = 15, 6, 6 for mild (green), moderate (orange), and severe (red) respectively in the early phase, and n = 18, 15, 17 for mild, moderate and severe in the late phase. (c) UMAP projections of live, CD45+ CD3+ cells from samples between D11-17 DPSO (equal numbers of cells sampled from n=7, 7, 7, 7 HD, mild, moderate, and severe patients, respectively) with FlowSOM clusters overlayed. (d) Marker expression in each FlowSOM cluster shown. (e) The percentage of each cluster derived from HD (blue), mild (green), moderate (orange) or severe (red) patients. (f) PBMCs were stimulated with peptides from the S, M, or N proteins in separate wells for 18 hours and IFN-γ production measured by ELISPOT. Response to S, M, and N peptides was summed and normalized per 100,000 cells plated. IFN-γ ELISPOT response shown as a linear correlation with DPSO with disease severity indicated by color with shaded area representing 95% confidence interval (top panel) and as a boxplot by disease severity (bottom panel); n = 10, 17, 18 in mild (green), moderate (orange), and severe (red), respectively. (g) Cytokine production after stimulation with pooled peptides from the S, M and N proteins, αCD3/CD28/CD49d antibodies, or PMA and ionomycin is shown for each individual patient. Background activity in unstimulated wells was subtracted from stimulated wells. The percentage of cells producing various combinations of IFN-γ, TNFα, and IL-2 were reported. HD: n = 9 for SMN stimulation, n = 8 for αCD3/CD28/ CD49d and PMA+ionomycin stimulation; mild n = 8; severe n = 7. (h) Maximum cytokine and chemokine levels related to T cell homeostasis are shown during the early phase (days 1-9 from symptom onset) and late phase (days 10-30 from symptom onset) of disease in SARS-CoV-2 infection compared to non-infected healthy controls. Each dot represents maximum value per individual subject during each phase of disease (early phase: mild, n=15; moderate, n=10; severe, n= 11 and late phase: mild, n=23; moderate, n=16; severe, n= 19 and non-infected healthy controls, n=18). (a, b, f, h) Significance was determined by two-sided Mann Whitney Wilcoxon test and p-values are indicated by asterisks (*, p ≤ 0.05; **, p ≤ 0.01; ***, p ≤ 0.001, ****, p ≤ 0.0001).

**Supplemental Fig 5:**
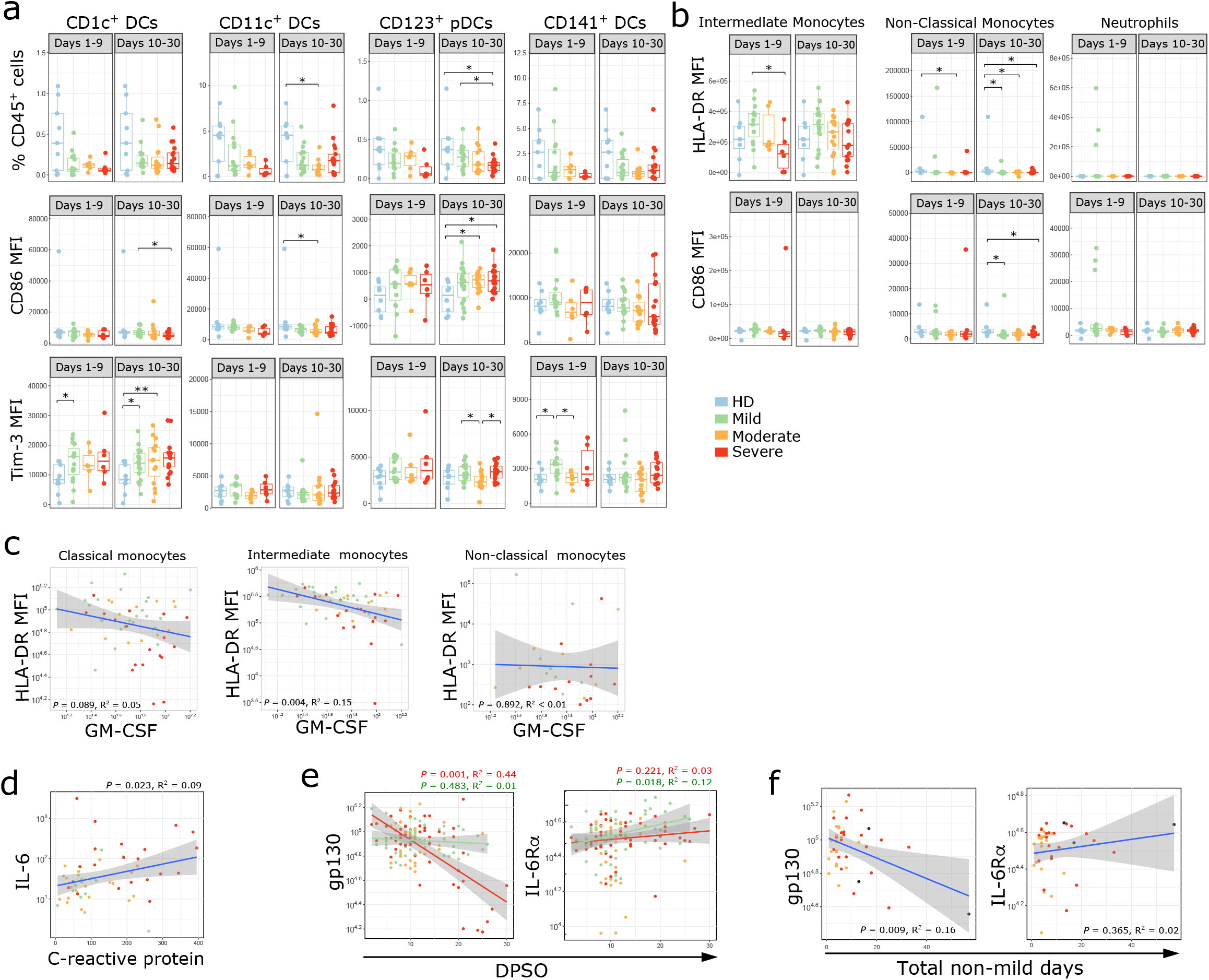
Induction of innate immune responses in SARS-CoV-2 infection. (a,b) Immune cell subsets related to the innate immune response are shown. Boxplots show percentages of each cell population out of live CD45+ cells or MFI of indicated markers on dendritic cells, monocytes or neutrophils. Where multiple timepoints within the early (D1-9) or late (D10-30) phase per patient were available, the mean was taken and each patient is represented by one dot per time phase. n = 9 for HD, n = 15, 6, 6 for mild, moderate, and severe respectively in the early phase, and n = 18, 15, 17 for mild, moderate and severe in the late phase. (c) The MFI of HLA-DR on classical, intermediate, and non-classical monocytes is shown as a linear regression with peak serum GM-CSF levels in n = 24, 13, 18 mild (green), moderate (orange), and severe (red) patients respectively. (d) Linear regression is shown for peak serum IL-6 and peak clinical CRP level. Each dot represents the maximum value per individual subject during the disease (mild, n=17; moderate, n=18; severe, n= 21). (e) Linear correlations between peak serum gp130 or Il-6R**α** levels versus DPSO in severe (red) and mild (green) patients; disease severity is indicated by color in mild (green, n = 23), moderate (orange, n= 19), and severe (red, n= 33) patients. (f) Peak individual serum levels of gp130 and IL-6R**α** are shown as linear correlations with the sum of days each patient spent in moderate or severe S/F status, termed “non-mild days”. Maximal disease severity indicated by color (moderate [orange, n=20]; severe [red, n= 19], or deceased [black, n= 3]). (a, b) Significance was determined by two-side Mann Whitney Wilcoxon test and p-values are indicated by asterisks (*, p ≤ 0.05; **, p ≤ 0.01; ***, p ≤ 0.001, ****, p ≤0.0001). (c-f) Shaded areas represent 95% confidence interval.

**Supplemental Fig 6:**
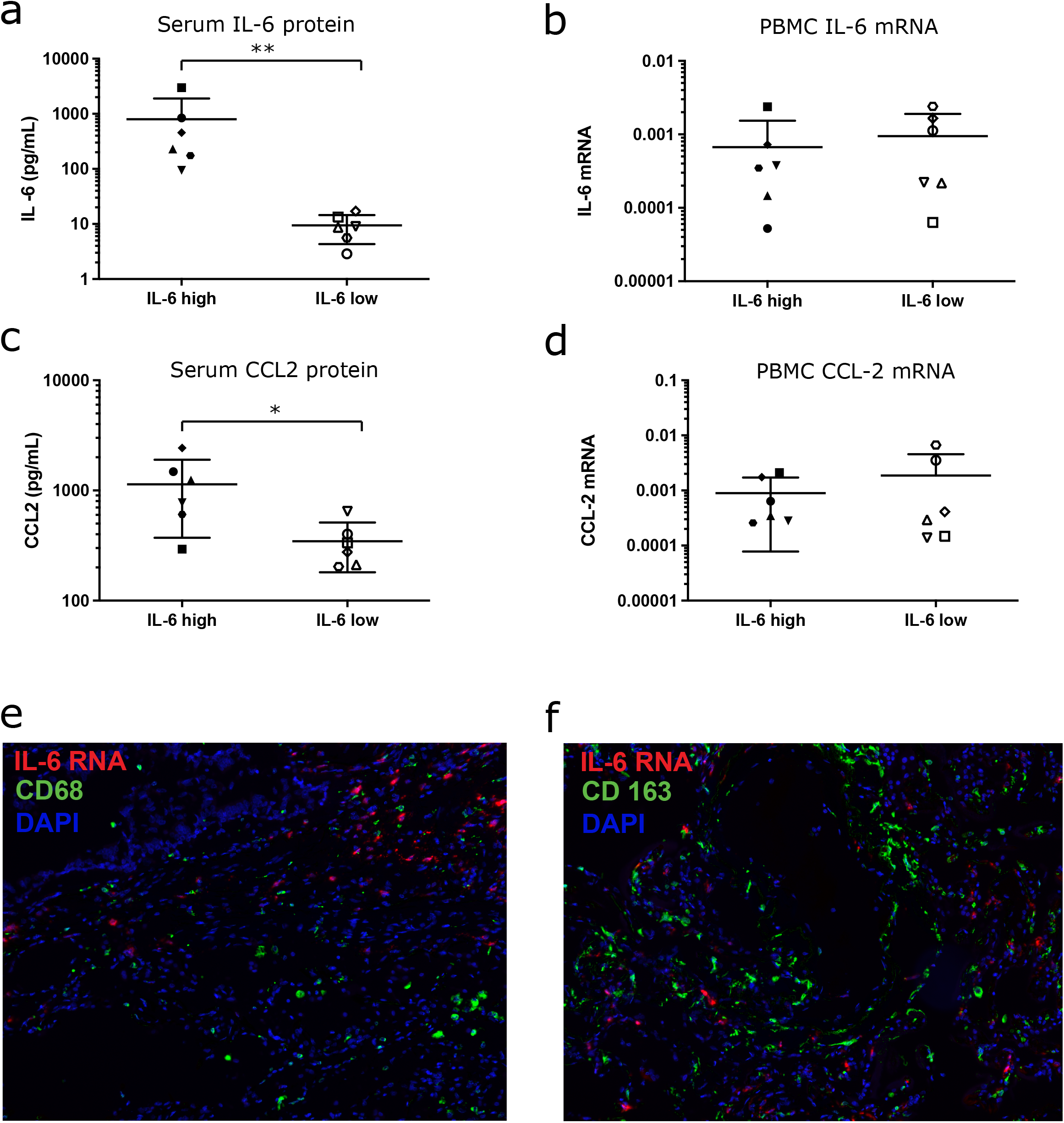
IL-6 and CCL2 are not predominantly produced by macrophages. (a-d) RNA was extracted from PBMCs from 6 patients who had either high or low levels of serum IL-6. Serum protein levels of IL-6 (a) and CCL-2 (b) in this cohort are shown. Expression of IL-6 (c) and CCL2 (d) mRNA from PBMCs was evaluated by RT-PCR. Error bars in (a-d) show mean + SD. Significance was determined by two-side Mann Whitney test and p-values are indicated by asterisks (*, p ≤ 0.05; **, p ≤ 0.01). (e-f) Representative image showing IL-6 expression outside of CD68+ (e) and CD163+ (f) macrophages. Multispectral images were acquired at 40x magnification. Multiplex immunofluorescence staining was performed for IL-6 RNA (red) and CD68 (panel e, green) or CD168 (panel f, green), and nuclear DAPI counterstain (blue).

**Supplemental Fig 7:**
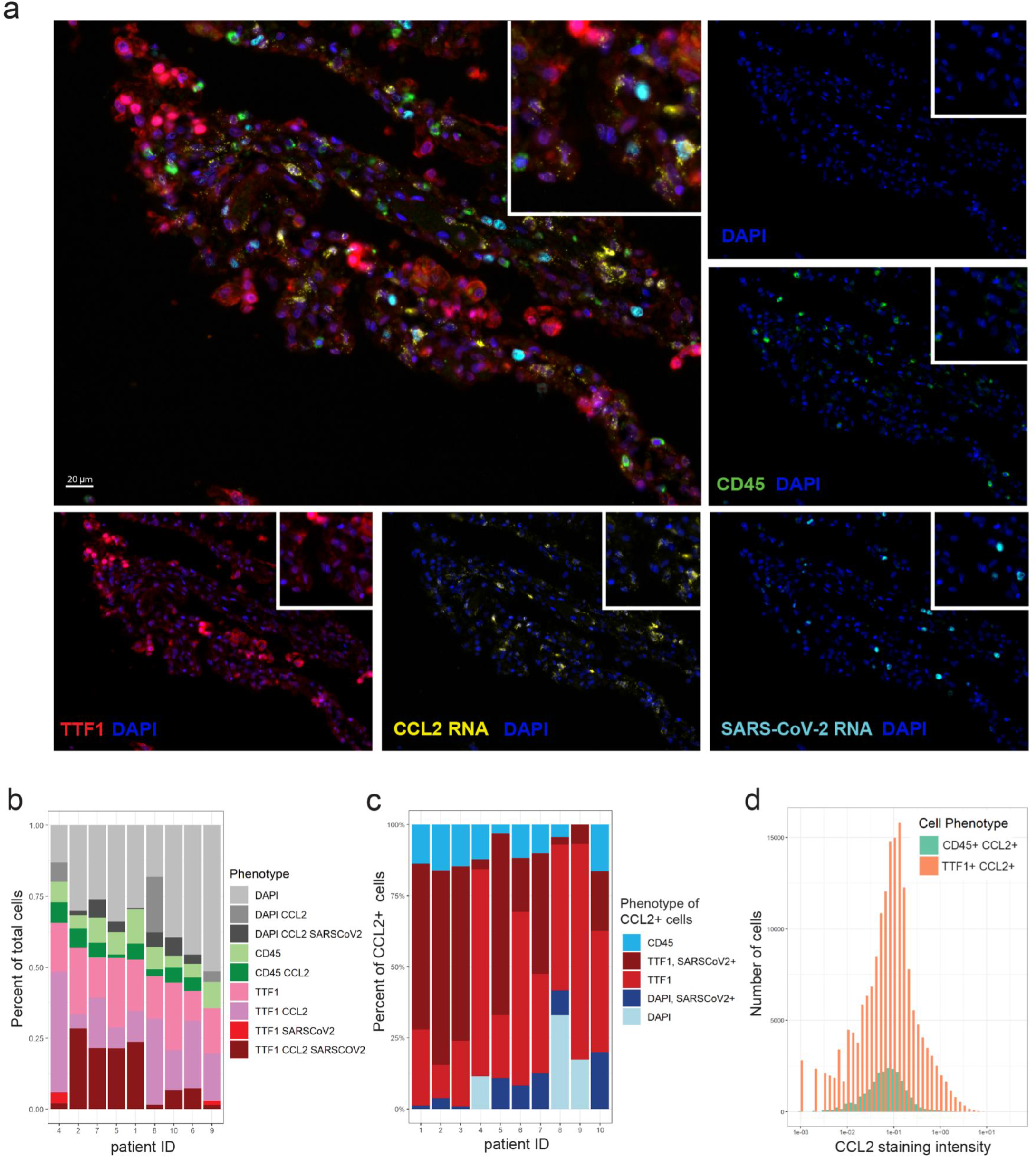
Cellular source of CCL2 in autopsy lung tissue in fatal COVID-19. (a) Representative staining for TTF1 (red), CD45 (green), CCL2 RNA (yellow), SARS-CoV-2 RNA (light blue), and nuclear DAPI counterstain (blue); each stain shown separately and merged. Overlaying high-power images showing TTF1+ pneumocytes expressing high levels of CCL2. (b) Bar plots showing the phenotype composition of cell populations in each autopsy lung specimen. (c) Bar plots showing the phenotype composition of CCL2+ cells in each autopsy lung specimen. (d) Histogram displaying the frequency distribution of mean staining intensity for CCL2 between TTF1+ CCL2+ cells (red) versus CD45+ CCL2+ cells (aqua). Cumulative data from all patients shown.

